# Comparative Effectiveness of Medical Concept Embedding for Feature Engineering in Phenotyping

**DOI:** 10.1101/2020.07.14.20151274

**Authors:** Junghwan Lee, Cong Liu, Jae Hyun Kim, Alex Butler, Ning Shang, Chao Pang, Karthik Natarajan, Patrick Ryan, Casey Ta, Chunhua Weng

## Abstract

**Objective:** Feature engineering is a major bottleneck in phenotyping. Properly learned medical concept embeddings (MCEs) have the semantics of medical concepts, thus useful for retrieving relevant medical features in phenotyping tasks. We compared the effectiveness of MCEs learned from knowledge graphs and electronic healthcare records (EHR) data in retrieving relevant medical features for phenotyping tasks.

**Materials and Methods:** We implemented five embedding methods including node2vec, singular value decomposition (SVD), LINE, skip-gram, and GloVe with two data sources: (1) knowledge-graphs obtained from the Observational Medical Outcomes Partnership (OMOP) common data model; and (2) patient level data obtained from the OMOP compatible electronic health records (EHR) from Columbia University Irving Medical Center (CUIMC). We used phenotypes with their relevant concepts developed and validated by the Electronic Medical Records and Genomics (eMERGE) network to evaluate the performance of learned MCEs in retrieving phenotype-relevant concepts. *Hits@k%* in retrieving phenotype-relevant concepts based on a single and multiple seed concept(s) was used to evaluate MCEs.

**Results:** Among all MCEs, MCEs learned by using node2vec with knowledge graphs showed the best performance. Of MCEs based on knowledge graphs and EHR data, MCEs learned by using node2vec with knowledge graphs and MCEs learned by using GloVe with EHR outperforms other MCEs respectively.

**Conclusion:** Medical concept embedding enables scalable feature engineering tasks, thereby facilitating high-throughput phenotyping. Knowledge graphs constructed by hierarchical relationships among medical concepts learn more effective MCEs, highlighting the need of more sophisticated use of big data to leverage EHR for phenotyping.

## INTRODUCTION

Phenotyping is a task of identifying a patient cohort underlying specific clinical characteristics. With the widespread adoption of electronic health records (EHR), phenotyping is one of the most fundamental research challenges encountered when using the EHR data for clinical research^1^. As learned from the Electronic Medical Records and Genomics (eMERGE) network, the process of developing and validating a phenotype requires a large amount of manual effort and time, typically up to 6-10□months^2,3^. A phenotype typically contains thousands to tens of thousands of relevant medical concepts. For example, the Type 2 Diabetes Mellitus (T2DM) phenotype developed by the eMERGE network contains about 12,000 relevant medical concepts^4,5^. Thus, identifying phenotype-relevant medical concepts (e.g., diagnosis, laboratory test, medication, and procedure concepts), which is called feature engineering, is essential but often labor-intensive step in developing a phenotype. Additionally, feature engineering in rule-based phenotyping largely relies on domain experts and can be error-prone and not generalizable or portable. Recently, data-driven high-throughput phenotyping methods have been proposed to readily extract relevant features from external knowledge sources^6-10^. Existing methods, however, often required text mining techniques that are not easy to implement, thus making it difficult to implement in real-world phenotyping tasks.

Medical concept embedding, which learns low-dimensional representations of medical concepts using embedding (i.e. neural embedding), has been used to improve the performance of various medical prediction tasks^11,12^ such as patient visit prediction^13^, patient outcome and risk prediction^14^, and phenotyping^15^ since properly learned MCEs capture the underlying semantics of medical concepts. Those MCEs are usually trained in unsupervised manner and fine-tuned with each downstream task.

Knowledge graphs are one of the widely used resources for learning MCEs. A knowledge graph contains medical concepts as nodes, which are connected via various relationships defined according to domain knowledge. Common knowledge graphs include Unified Medical Language System (UMLS), Systematized Nomenclature of Medicine Clinical Terms (SNOMED CT), International Classification of Disease (ICD), and Human Phenotype Ontology (HPO). Graph-based embedding methods have been leveraged to capture the diversity of connectivity patterns observed in graphs to learn the node embedding for medical concepts^16^. For example, Agarwal et al. learned embedding of medical concepts that showed impressive performance in various healthcare application tasks including multi-label classification and link prediction from SNOMED CT using various graph embedding methods^17^. A knowledge graph can be enriched by introducing other kinds of relationships to increase connectivity of nodes in the knowledge graph as Shen et al. learned efficient embeddings for HPO concepts by enriching the HPO knowledge graph leveraging heterogeneous vocabulary resources^18^.

EHR are another popular resource to learn MCEs. A patient visit triggers a number of medical concepts being documented in the EHR. A typical strategy to utilize EHR for learning MCEs is to consider each visit or pre-determined time window in a patient EHR as a bag of medical concepts. Embedding methods that leverage co-occurrence information, such as GloVe^19^ and skip-gram^20^, can then be adopted to learn MCEs. For example, Choi et al. developed Med2vec, which leveraged patient visits to learn MCE and showed strong performance in patient’s future visit and status prediction^13^.

We posit that properly learned MCEs have the potential to provide a scalable approach to retrieve phenotype-relevant concepts, thereby mitigating feature engineering efforts of phenotyping and accelerating phenotype development. In this study, we compared how effectively MCEs learned from various data sources can facilitate feature engineering for phenotyping. We trained MCEs using five different embedding methods with two different data sources – knowledge graphs and patient level data obtained from EHR. The phenotypes validated by the eMERGE network^2^ with their corresponding medical concept lists were used to evaluate different MCEs. MCEs were assessed on retrieving phenotype-relevant concepts given seed concepts extracted from each phenotype. Additionally, we also provided the learned MCEs in a concept recommender system through web application programming interface (API), which can be easily implemented.

## MATERIALS AND METHODS

### Data Description and Processing

All concepts used in this study are based on the Observational Health Data Science and Informatics (OHDSI) Observational Medical Outcomes Partnership common data model (OMOP CDM). OHDSI is a multi-stakeholder, interdisciplinary collaborative that aims to bring out the value of health data through large-scale analytics^21^. The OMOP CDM harmonizes several different medical coding systems, including but not limited to ICD-9-CM, ICD-10-CM, SNOMED CT, and LOINC, to achieve standardized vocabularies while minimizing information loss, thus provides a unifying data format for various analysis pipelines^22^. The standard vocabularies were built by OMOP CDM and defined the meaning of a clinical entity uniquely across all databases and independent from the coding system. Non-standard concepts that have the equivalent meaning to the standard concept were then mapped to the standard concept. We focus on condition (i.e. diagnosis) concepts in this study, which play a critical role in phenotyping.

We used two kinds of knowledge graphs in this study, hierarchical knowledge graph and enriched knowledge graph. The hierarchical knowledge graph was constructed by using *concept_relationship* table, which contains “is-a” and “subsume” relationships between standard condition concepts. The enriched knowledge graph expands upon the hierarchical knowledge graph by adding non-standard concepts connected to standard concepts by hierarchical relationship from the *concept_ancestor* table, thus increasing the connectivity of the knowledge graph and expanding the number of concept nodes in the graph. An example of hierarchical knowledge graph and enriched knowledge graph can be found in **Supplementary Figure 1**. Summary statistics of the knowledge graphs are provided in **Table 1**.

**Table 1.** Summary statistics of the knowledge graphs.

|  | **Hierarchical**<br>**knowledge graph** | **Enriched**<br>**knowledge graph** |
| --- | --- | --- |
| # of unique concepts (concept nodes) | 306,266 | 312,089 |
| # of edges (unique relationships) | 588,298 | 671,591 |
| Avg. # of edges per concept node | 1.93 | 2.15 |

EHR data were obtained from the Columbia University Irving Medical Center (CUIMC) EHR clinical data warehouse containing inpatient and outpatient data starting from 1985. The CUIMC EHR data has been converted to the OMOP CDM and covers more than 36,000 medical concepts from multiple domains (e.g., condition, drug, and procedure) extracted from more than 5 million patients. To ensure data quality, we used the recent 5-year EHR data from 2013 to 2017^23^. We processed the EHR data into the format of bag-of-medical concepts by applying visit window and 5-year window on each patient’s EHR. **Figure 1** depicts how the 5-year and visit window were applied to the EHR. We excluded windowed-EHR containing only a single concept since they do not provide any meaningful co-occurrence information. Summary statistics of the EHR are provided in **Table 2**.

**Table 2.** Summary statistics of windowed EHR. Windowed-EHRs containing only a single concept were excluded.

|  | **visit-windowed EHR** | **5-year-windowed EHR** |
| --- | --- | --- |
| # of patients | 1,292,369 | 1,370,787 |
| # of windowed records | 8,865,691 | 1,370,787 |
| # of unique concepts | 17,175 | 17,288 |
| Avg. (std) # of concepts<br>per windowed record | 4.3 (3.3) | 12.4 (17.2) |

**Figure 1.**
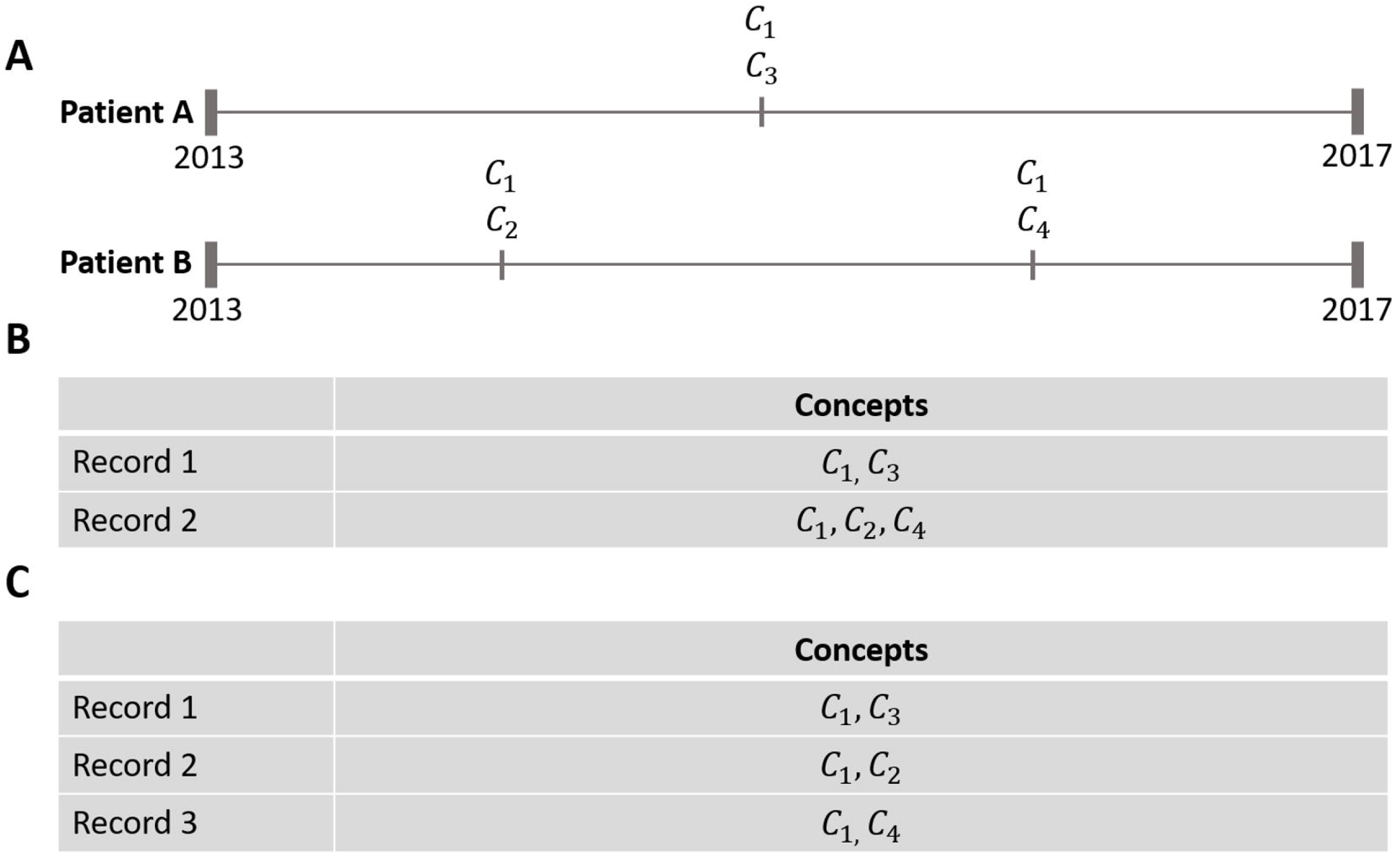
Application of visit window and 5-year window to patient records. In (**a**), Each mark along a patient’s timeline indicates a visit containing medical concept(s), *C*_*x*_. Raw patient records (**a)** are transformed to the formats in (**b**) by applying a 5-year window and in (**c**) by applying visit windows. Visits containing only a single medical concept were excluded. For example, the last visit of the patient A in (**a**) was excluded since the visit only contains a single concept.

### Learning Medical Concept Embedding from EHR Data

#### GloVe

GloVe was originally developed in the natural language processing domain for learning word representations, which uses the global co-occurrence statistics of words in a corpus to learn the word representations^19^. Although GloVe was designed to learn word representations, by treating medical concepts as words and windowed-EHR as a bag-of-concepts, we can leverage GloVe to learn the representations of medical concepts. *GloVeEmb_V* and *GloVeEmb_5Y* were trained by implementing GloVe on co-occurrence statistics of the visit windowed EHR and the 5-year windowed EHR respectively. Both are trained for 30 epochs and the maximum co-occurrence was set to 100.

#### Skip-gram

Skip-gram learns the representations of words considering their local neighborhood in the given context window^20^. *SGEmb_V* and *SGEmb_5Y* were trained for 30 epochs by implementing skip-gram on the visit and 5-year windowed EHR respectively.

### Learning Medical Concept Embedding from Knowledge Graphs

#### Singular value decomposition (SVD)

Singular value decomposition (SVD) is one of the commonly used traditional matrix factorization methods, which factorizes a data matrix into lower dimensional matrices. We obtained low-dimensional representation for the concepts (i.e. embedding) using SVD on the adjacency matrix of the knowledge-graphs. In this study, *SVD* and *SVD+* were obtained by implementing SVD on the hierarchical and enriched knowledge graph respectively.

#### node2vec

node2vec^24^ learns embedding for the nodes in a graph using random walk. The embedding of the nodes learned by node2vec has information regarding homophily and structural equivalence of the nodes by adopting two random walk parameters that govern breadth-first search and depth-first search. *n2vEmb* and *n2vEmb+* were trained by implementing node2vec on the hierarchical and enriched knowledge graph for a single epoch respectively. Hyperparameters were set as provided in the original publication^24^: walk-length=10; window-size=10; in-out parameter (*q*)=1.0; return parameter (*p*)=1.0.

#### Large-scale information network embedding (LINE)

Large-scale information network embedding (LINE) learns embedding for the nodes in a graph by approximating first-order proximity and second-order proximity^25^. *LINEEmb* and *LINEEmb+* were trained by implementing LINE on the hierarchical and enriched knowledge graph for five epochs respectively. The negative ratio was set to five described as the default setting in the original publication^25^.

### Implementation Details

All MCEs were trained on a machine equipped with 2 × Intel Xeon Silver 4110 CPUs, 188GB RAM, and one Nvidia GeForce RTX 2080 TI GPU. We implemented SVD using Numpy 1.18.5 on Python 3.7.1. GloVe and skip-gram were implemented using Tensorflow 2.2.0^26^ on Python 3.7.1. LINE and node2vec were implemented using OpenNE (available at https://github.com/thunlp/OpenNE), a python package for network embedding. The source codes are publicly available at https://github.com/WengLab-InformaticsResearch/mcephe. The concept recommender API is provided at https://github.com/WengLab-InformaticsResearch/concept-recommeder.

### Evaluation Strategy

To assess the performance of learned MCEs in retrieving relevant medical concepts for phenotypes, we used 33 independently validated phenotyping algorithms from the eMERGE network with their corresponding code books. The eMERGE network created the Phenotype Knowledgebase (PheKB) to facilitate phenotyping and sharing of the phenotyping knowledge, where phenotypes are shared as descriptive text, workflow charts, and code books of medical concepts (available at https://www.phekb.org/). The Columbia eMERGE team converted PheKB code books by implementing the OMOP CDM, thus we obtained 20,640 condition concepts in 33 phenotypes. We excluded the concepts related to phenotype exclusion criteria. The number of concepts in each phenotype is provided in **Supplementary Table 1**.

Since the number of unique trainable concepts trained in each of the MCEs is different from each other, we constructed a standard evaluation set for each MCE to create a comparable evaluation. The standard evaluation set consists of the intersection between trainable concepts in the respective MCE and concepts from all phenotypes in PheKB (**Figure 2**). We excluded the out-of-bag concepts (i.e. concepts from PheKB phenotypes but not used to train the MCEs and vice versa) from the standard evaluation set. For example, if concept *C* appears in the knowledge graph but never appears in any of 33 phenotypes in PheKB, concept *C* was excluded from the standard evaluation set with regard to the knowledge graph since it is impossible to evaluate the embedding of the concept *C*.

**Figure 2.**
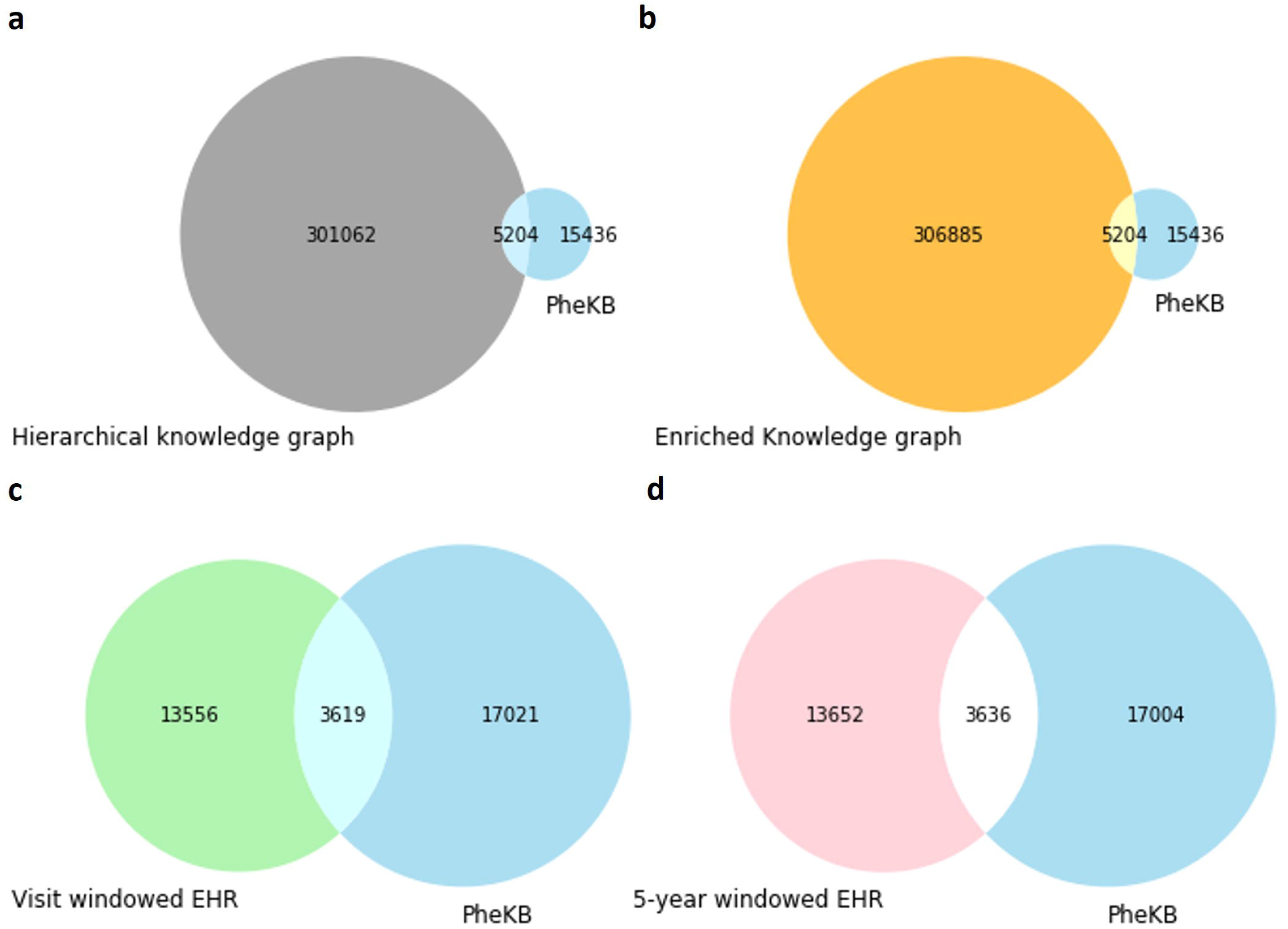
Set diagrams between the unique concepts in PheKB and in each medical concept embedding (MCE). The intersection of each set diagram is the standard evaluation set for that MCE. Since we excluded EHRs that had less than two concepts in each window, there are slight difference in the total number of unique concepts between visit windowed and 5-year windowed EHR.

In real-world applications, feature engineering for phenotyping is often started by generating seed concepts or features. Thus, we quantitatively assessed different MCEs’ performance in retrieving phenotype-relevant concepts given varying number of seed concepts, from a single seed concept to multiple seed concepts. We also assessed the interpretability of learned MCEs by plotting on 2-dimensional space.

#### Evaluation based on a single seed concept

*Hits@k* is a commonly used metric to assess how well the retrieved results satisfy a user’s query intent in information retrieval. In our evaluation, the query was the seed concept(s), and the retrieved results were candidate concepts. Given a single seed concept selected from a phenotype, each MCE retrieved the candidate concepts nearest to the seed concept based on the embedding distance. We used cosine similarity to measure the distance between embeddings. Since the number of relevant concepts in a phenotype varies across the 33 phenotypes, we used a modified version of *Hits@k* – *Hits@k%* as our evaluation metric to provide a more consistent comparison across different phenotypes. Specifically, *Hits@k%* for phenotype *p* based on MCE *e* is defined as Eq (1):

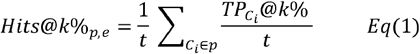

where *t* is the number of unique concepts in phenotype *p*, 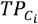 is the number of relevant concepts retrieved for the embedding of the seed concept *C*_*i*_, and *k%* is the percentage that controls the number of nearest concepts retrieved for the seed concept based on the cosine similarity for each phenotype. We reported the average *Hits@k%* for each MCE by averaging the results from all phenotypes.

#### Evaluation based on multiple seed concepts

We again evaluated the *Hits@k%*, except this time using multiple concepts selected from a phenotype for the seed concepts. Given *n* seed concepts derived from a phenotype, the embedding for the seed was obtained by summing the embeddings of all *n* seed concepts, then each MCE retrieved the candidate concepts nearest to the seed embedding. Seed concepts for a phenotype were randomly chosen from the set of concepts in the phenotype. The number of seed embeddings was set to the number of unique concepts in the phenotype, yielding the same number of seed embeddings as in the single concept seed case. We performed this evaluation with *n* = 5. Similar to before, *Hits@k%* for phenotype *p* based on MCE *e* are defined as Eq (2):

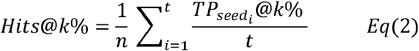

where 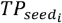 is the number of relevant concepts retrieved for the embedding of the *seed*_*i*_. Average *Hits@k%* was reported for each MCE.

#### Visualization of learned MCEs

To assess interpretability of the learned embeddings, we plotted the embeddings of 1,221 concepts, which lies in the intersection between the evaluation set of all MCEs, on 2-dimensional space using t-SNE^27^. For clear visualization, we excluded concepts that were included in multiple phenotypes in the process of selection of concepts, which made 26 phenotypes left.

## EXPERIMENT RESULTS

### Overall Performance

**Figure 3** shows the average *Hits@k%* of all MCEs based on a single seed concept and five seed concepts. Of all MCEs based on both single seed concept and multiple seed concepts scenarios, *n2vEmb+* outperforms all other MCEs in average *Hits@k%*. MCEs learned by using node2vec shows better performance than other knowledge graph-based MCEs. Among EHR-based MCEs, MCEs learned by using GloVe showed better performance than MCEs learned by using skip-gram.

**Figure 3.**
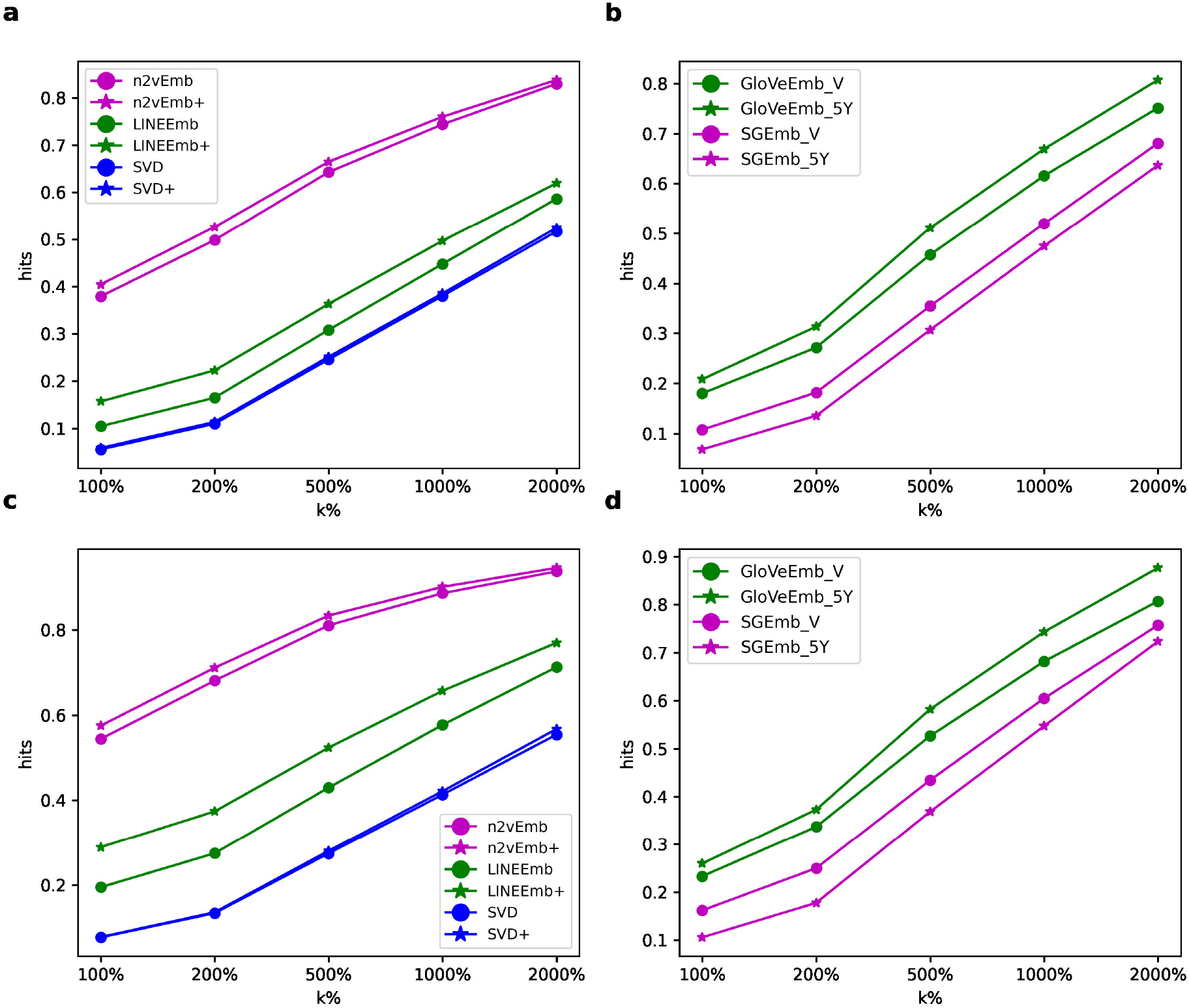
*Hits@k%* of the embeddings learned from (**a, c**) knowledge graphs and (**b, d**) EHR data based on a single seed concept and five seed concepts respectively.

### Performance on Individual Phenotypes

**Figure 4** shows average *Hits@500%* of all phenotypes for each MCE based on a single seed concept (**4a**) and five seed concepts (**4b**). *Hits@k%* for all the other *k%* based on a single and five seed concept(s) are provided in **Supplementary Table 2**-**21**. We excluded one phenotype (*Diverticulosis*), which contains less than 10 concepts, from the evaluation based on five seed concepts. In both evaluations based on a single concept seed and on five seed concepts, *n2vEmb+* had the best performance among the MCEs in more than half of the evaluated phenotypes.

**Figure 4.**
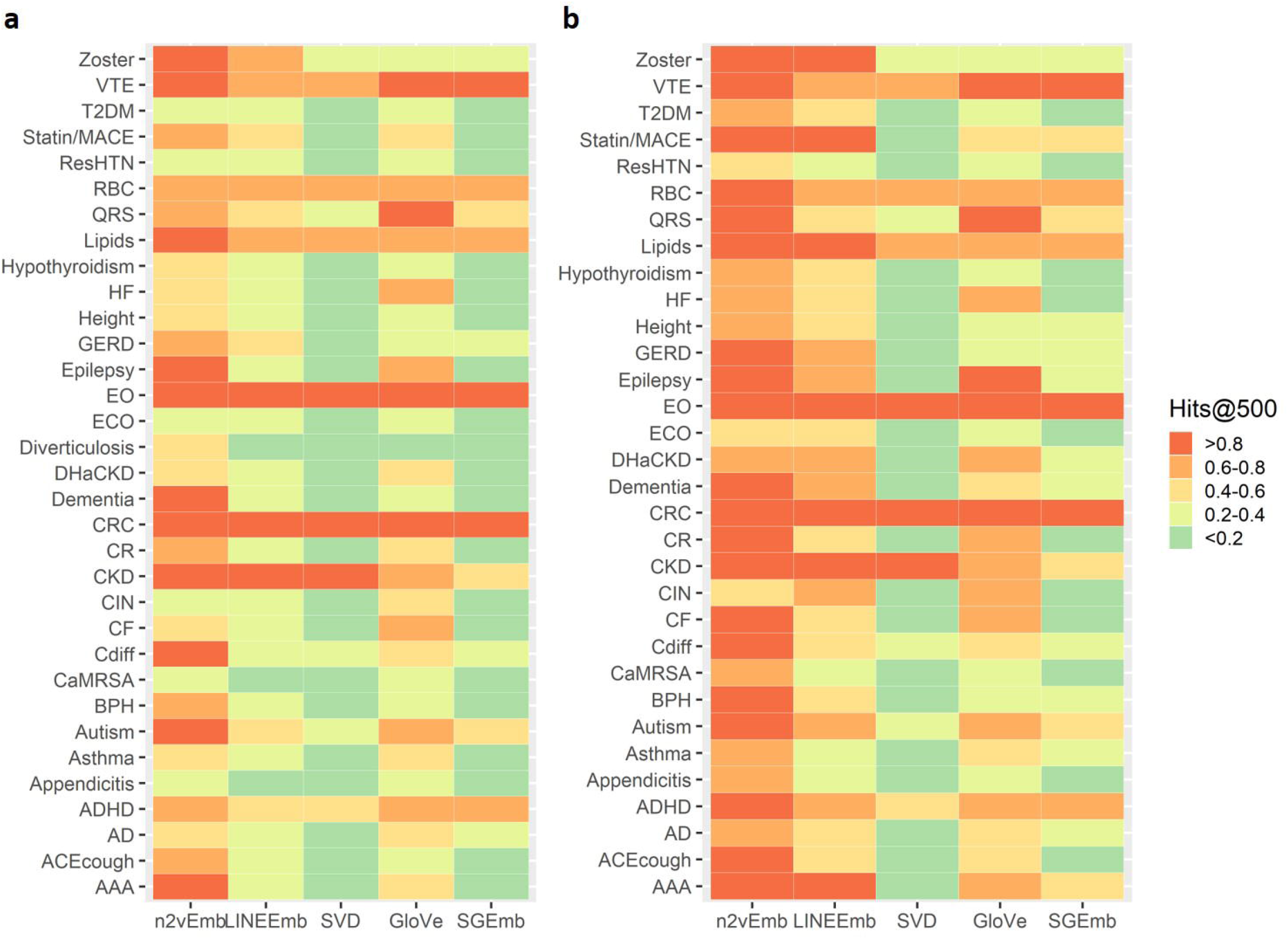
Average *Hits@500%* of all individual phenotypes based on (**a**) a single seed concept and (**b**) five seed concepts for MCEs. Full names for abbreviated phenotypes are provided in **Supplementary Table 1**.

### Visualization of Learned MCEs

t-SNE scatterplots of the 1,221 concepts for all MCEs are shown in **Figure 5**. The color of each marker represents the phenotype that the concept belongs to.

**Figure 5.**
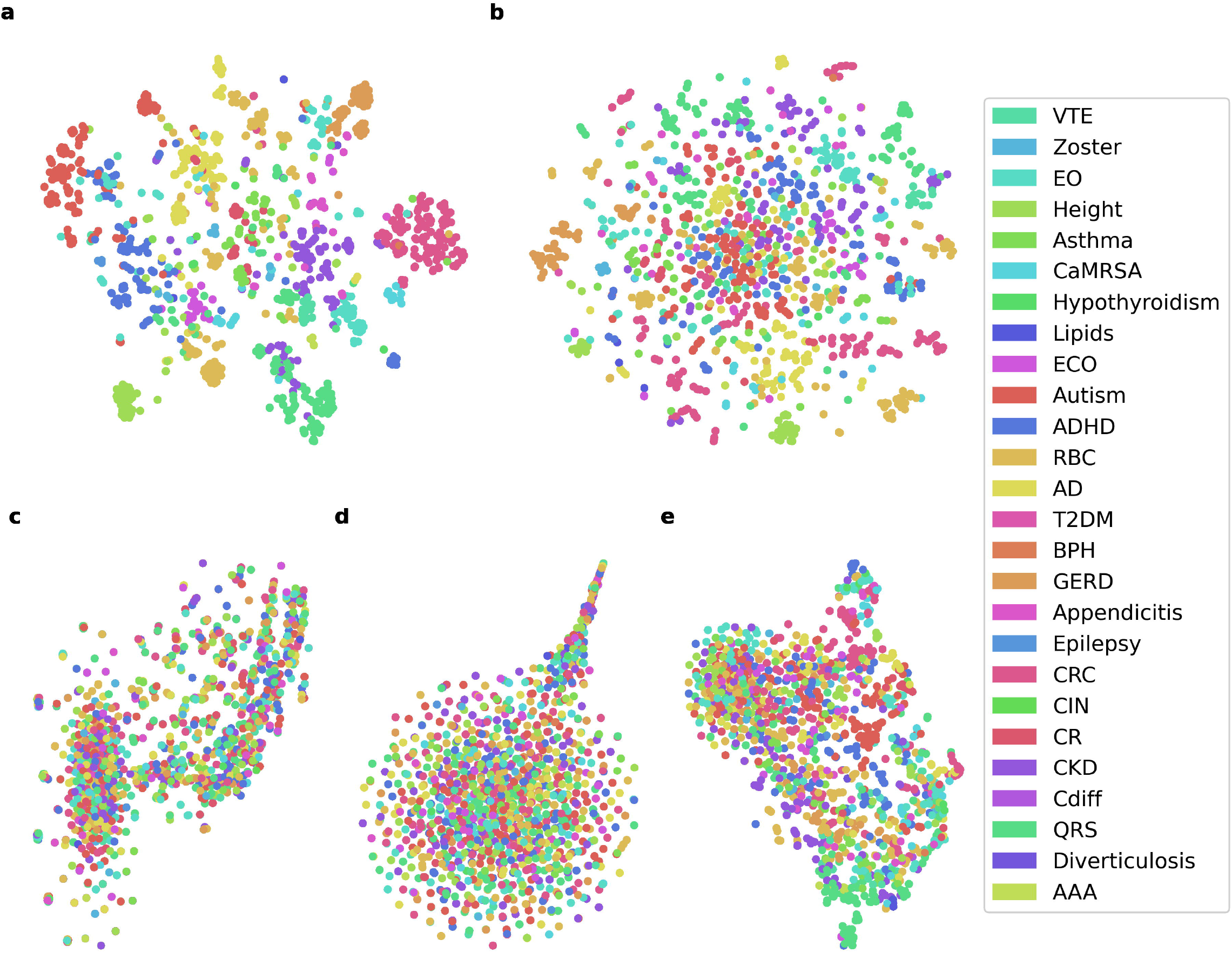
t-SNE scatterplots of the 1,221 concepts, which lies in the intersection between the evaluation set of all MCEs, for (**a**) *n2vEmb+*, (**b**) *LINEmb+*, (**c**) *SVD+*, (**d**) *SGEmb_5Y*, and (**e**) *GloVeEmb_5Y*. Since we excluded concepts that were included in multiple phenotypes in the process of selection of concepts, there were only 26 phenotypes in the scatterplots. Full names for abbreviated phenotypes are provided in **Supplementary Table 1**.

### Impact of Hyperparameters

Hyperparameters of unsupervised learning methods are often optimized and fine-tuned with a specific downstream task. Besides using the default settings of hyperparameters suggested from original publications of the methods to assess MCEs, we investigated the impact of important hyperparameters for each method. **Table 3** lists some important hyperparameters for each method. **Figure 6** shows the impact of the hyperparameter on the performance for each MCE. Hyperparameters of each method except the controlled hyperparameter were set to the default setting as described in each method section above. Training epochs of *GloVeEmb_V* and *SGEmb_V* were set to 10 to reduce time for training. We did not experiment the different context window of skip-gram since the impact of varying size of the window was already experimented in **Figure 3**.

**Table 3.** Some important hyperparameters for each embedding method.

| Embedding method | Hyperparameters |
| --- | --- |
| GloVe | - $x_{max}$ : the maximum number of co-occurrences so that frequent co-occurrences are not overweighted;<br>- Embedding dimensionality |
| skip-gram | - Context window: the size of window to be considered as neighborhood for the target concept<br>- Embedding dimensionality |
| SVD | - Embedding dimensionality |
| node2vec | - Return parameter ($p$): a parameter that controls the likelihood of immediately revisiting a node in the random walk. High $p$ value ensures the random walk to less likely sample an already visited node in the following two steps;<br>- In-out parameter ($q$): a parameter that controls the degree of breadth-first search (BFS) and depth-first search (DFS). High $q$ value makes the random walk more inclined to BFS.<br>- Embedding dimensionality |
| LINE | - Epochs: The number of training epochs. LINE optimally requires different number of epochs, which is proportional to the size of the input graph;<br>- Embedding dimensionality |

**Figure 6.**
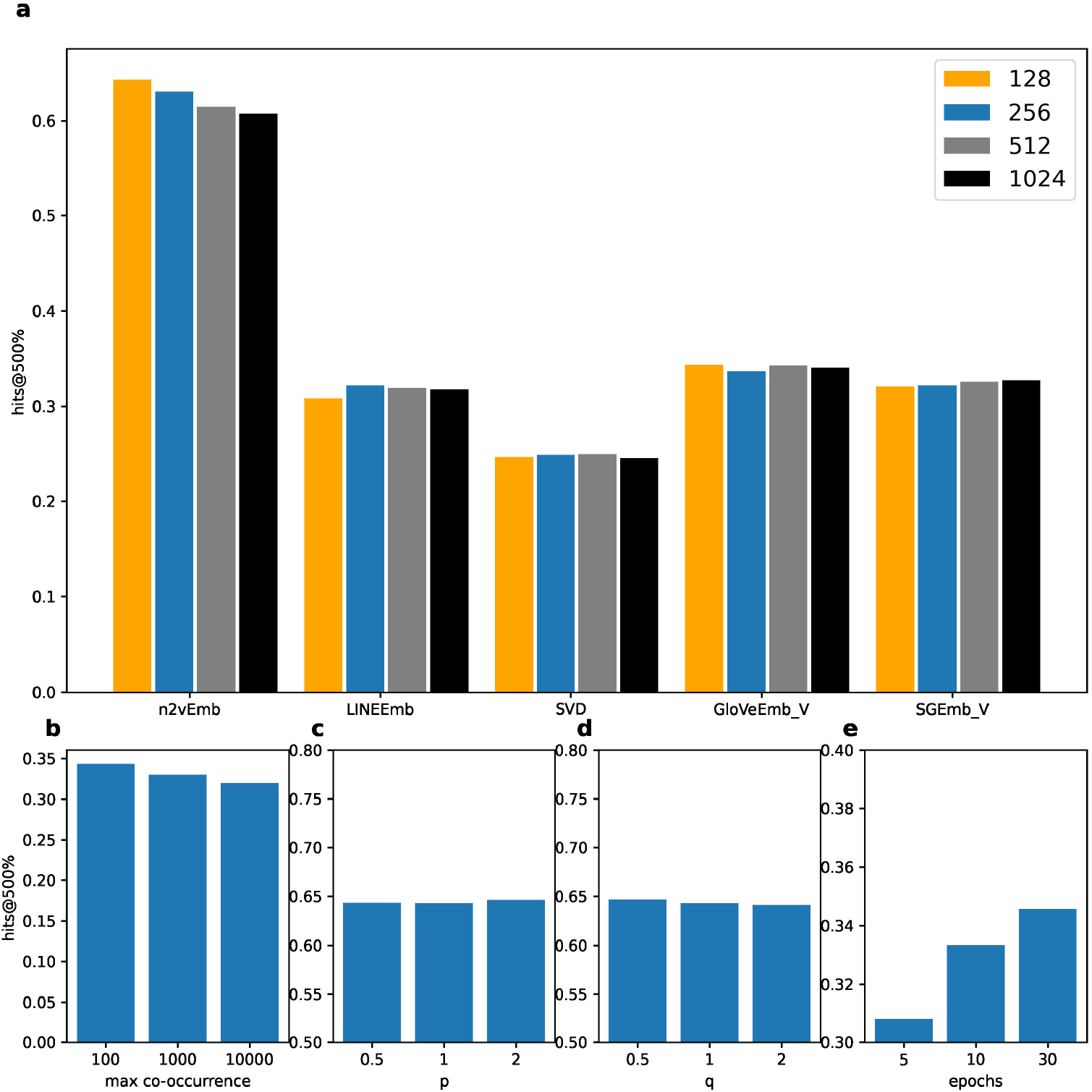
The impact of the hyperparameters on the performance for each MCE. (**a**) *Hits@500%* with different embedding dimension for *GloVeEmb_V, SGEmb_V, SVD, n2vEmb*, and *LINEEmb*. ***(b)****Hits@500%* with different *x*_*max*_ (maximum number of co-occurrences) of *GloVeEmb_V*. (**c** and **d**) *Hits@500%* with different *p* and *q* of *n2vEmb*. (**e**) *Hits@500%* with different number of epochs of *LINEEmb. GloVeEmb_V* and *SGEmb_V* were trained for 10 epochs.

## DISCUSSION

In this study, we assessed the potential of different MCEs for feature engineering in phenotyping. We evaluated MCEs learned by using five methods with two different data sources: knowledge graphs and EHR data. MCEs learned by using node2vec with knowledge graphs achieved the best performance in retrieving phenotype-relevant concepts. We provided a concept recommender system that can be used for feature engineering in phenotyping task leveraging the learned MCEs, which is easily implemented.

Among three different graph-based embedding methods, node2vec achieved the best performance. SVD showed the worst performance, which indicates phenotyping tasks often involve complex relationships among nodes that is more than first-order proximity that can be captured by simple matrix factorization. While LINE and node2vec both consider the first- and second-order proximity to learn the embeddings of the nodes, node2vec is able to reuse the samples via a random walk strategy. Our results suggest that node2vec learns better MCEs for feature engineering task in phenotyping, indicating random walk is an important strategy to learn more efficiently from the knowledge graphs having many nodes but not dense. We admit that there are other popular graph-based embedding methods except the methods we used in this study such as Graph Convolutional Networks (GCN) and DeepWalk. Since GCN currently cannot be implemented on a large graph and node2vec can be considered as a generalized version of DeepWalk (i.e. node2vec when *p*=1 and *q*=1), we did not include those methods in this study.

The difference of performance between MCEs based on the hierarchical knowledge graph and the enriched knowledge graph showed that enriching a knowledge graph by introducing additional relationships that have connection with the existing nodes in the knowledge graph is beneficial for learning the embedding of the nodes. This finding aligns well with the result from Shen et al., where the authors obtained efficient embedding for the concepts in HPO using an enriched knowledge graph^18^. It is not always true, however, that enriching a knowledge graph will lead to efficient embedding of the nodes. For example, introducing a singular node that is connected to less than one existing node cannot lead to efficient learning since the singular node does not increase connectivity of the knowledge graph, which is critical for learning efficient embedding of the nodes. This can hinder MCEs from leveraging a knowledge graph that is built upon concepts from multiple distinct domains but lack of inter-domain relationships. The current version of OMOP CDM also has this limitation, since there are only 22,334 relationships between condition and drug concepts compared to 2,148,636 and 23,435,796 relationships between condition-condition and drug-drug concept pairs, respectively, in the *concept_relationship* table.

Of EHR-based methods, GloVe achieved the best performance. The better performance of GloVe can be explained by its ability to learn by using global co-occurrence statistics instead local context windows. Besides GloVe and skip-gram used in this study, there are many other word embedding methods that can be leveraged to learn MCEs including but not limited to sequence-based embedding (e.g., ELMo) and attention-based embeddings (e.g., BERT). We did not include those models since there are no strict order of the medical concepts within each window of the patients. One possible future research question is to apply those state-of-the-art models to learn MCEs leveraging EHR.

Among *GloVeEmb_5Y* and *GloVeEmb_V*, 5-year window showed better performance than visit window. This is perhaps because phenotype-relevant condition concepts often appear across multiple visits and there are irregular time intervals between visits. For example, concepts related to heart failure including initial presenting signs, symptoms and complications appear in multiple visits with the progression of heart failure. The 5-year window, which covers multiple visits, can capture sematic relationships between concepts with long-term relationships better than the visit window. 5-year window also builds a less sparse co-occurrence matrix of the concepts than visit window, which could help capture the underlying semantic relationships between medical concepts. Among *SGEmb_5Y* and *SGEmb_V*, however, visit window showed better performance than 5-year window. The different impact of window size in using GloVe and skip-gram is because skip-gram simply tries to minimize the distance between the concepts co-occurred in the same window, while GloVe utilizes global co-occurrence statistics to cancel out the noise from non-discriminative concepts.

The window size of EHR can be adjusted with respect to the data quality, purpose of a study and method to be used. We decided to use CUIMC EHR data from 2013 to 2017 with visit and 5-year window to ensure data quality of the EHR^23^. A large window size, however, could introduce noise into the co-occurrence information for some acute diseases where intra-visit information between concepts are more important than inter-visit information. Therefore, if one aims to learn MCEs using EHR for feature engineering of a specific phenotype, careful consideration of the characteristics of the phenotype will be required while selecting the window size. For example, visit window can be used to learn MCEs for phenotyping acute diseases such as clostridioides difficile and a larger window (e.g., lifetime window or 5-year window) can be used to learn MCEs for phenotyping the diseases where symptoms appear in a long period of time such as heart failure and chronic kidney disease.

From **Figure 3**, we can see that all MCEs showed improved performance with the increasing number of seed concepts used to generate the seed embedding. However, as a trade-off for the improved performance, providing carefully selected seed concepts requires more effort from domain experts.

We can see from **Figure 5** that *n2vEmb+* and *LINEEmb+* showed better embeddings that align with phenotypes than other MCEs. From the figure, we observed more recognizable clusters that align with phenotypes in *n2vEmb+* and *LINEEmb+* than other MCEs. This result suggests that co-occurrence information from EHR may not be sufficient for learning interpretable MCEs that are consistent with phenotypes. It is interesting to see that other studies also found that simple co-occurrence information cannot learn interpretable representations that align with medical knowledge, although they did not use phenotyping knowledge to assess interpretability^28,29^. Nevertheless, co-occurrence information from EHR can reflect the daily clinical operation, providing complementary information to ontological knowledge for phenotype development.

From **Figure 6**, we can see the impact of important hyperparameters of each method on MCE’s performance. Most of the MCEs showed saturated performance with less than 256 dimensionality, while skip-gram showed slight increase of the performance with increasing dimensionality. For MCEs learned by using GloVe, low *x*_max_ resulted in better performance. For MCEs learned by using node2vec, better performance was achieved with larger *p* and smaller *q* perhaps because the graphs are not dense. For MCEs learned by using LINE, better performance was achieved with increasing number of epochs. It may seem natural that more epochs lead to better performance, however, this is not always true for LINE based on other studies^16^. Therefore, careful selection of the number of training epochs is required with consideration of the size of the graph.

There are several limitations in the study that should be noted. First, the dataset for evaluating MCEs was obtained from the phenotypes developed and validated by eMERGE network. Since most of the eMERGE phenotypes were developed by using rule-based algorithms, our findings in this study might not be generalized well to the phenotypes developed by machine learning based methods. Second, this study investigated the performance of the MCEs only using the condition (i.e. diagnosis) concepts. Considering that concepts from several different domains are often involved in phenotype development, future works will be required to investigate the performance of MCEs using concepts from other domains. Finally, representation learning including neural embedding is a rapidly evolving field. We admit that besides the embedding methods investigated in this study, there are other more sophisticated methods. Although this study focused on evaluation of MCEs learned by using knowledge graphs and EHR data, our evaluation framework can be generalized to wide range of MCEs learned by using diverse data sources and methods.

### Case Study of False Positive Concepts

Since the eMERGE phenotypes we used in the study were developed using rule-based algorithms, some of the false positive concepts retrieved by using MCEs might still be useful for feature engineering in phenotyping. We thus qualitatively investigated the false positive concepts in the retrieved concepts (i.e. retrieved as candidate concepts but not included in the concept list of eMERGE phenotype) for a selected phenotype - *Type 2 Diabetes Mellitus* (*T2DM*) based on *n2vEmb+* and *GloVeEmb_5Y*. We first manually selected five seed concepts and obtained top 50 retrieved concepts given the seed. The false positive concepts were scored based on whether the concept is relevant to the phenotype: 1 was assigned for the concept that is considered as strongly relevant to the phenotype and can directly be included in the phenotype; 0.5 was assigned for the concept that is considered as relevant to the phenotype and can be used a covariate concept or proxy concept for developing the phenotype; and 0 was assigned for the concept that is considered as irrelevant to the phenotype. The selection of the seed concepts and scoring of the false positive concepts were conducted with help of a clinically experienced researcher. **Table 4** showed the average relevant score of the false positive concepts for *T2DM* based on *n2vEmb+* and *GloVeEmb_5Y*. Among 27 and 40 false positive concepts for *n2vEmb+* and *GloVeEmb_5Y*, 67% and 50% of them were confirmed to be included in the phenotype directly or used as covariate concepts for feature engineering of the *T2DM* respectively. This result suggests that although there were some phenotypes that showed low *Hits@k%*, it does not necessarily mean the MCE is not useful for feature engineering tasks since *Hits@k%* was measured by using the eMERGE phenotypes as a gold standard.

**Table 4.** Average relevant score of the false positive concepts for *Type 2 Diabetes Mellitus* based on *n2vEmb+* and *GloVeEmb_5Y*.

|  | *n2vEmb+* | *GloVeEmb_5Y* |
| --- | --- | --- |
| # of false positive concepts in the 50 retrieved concepts | 27 | 40 |
| # of false positive concepts that can be used directly or used as covariate concepts | 18 | 20 |
| Average relevant score of the false positive concepts | 0.473 | 0.275 |

## CONCLUSIONS

We assessed the potential of several different MCEs for feature engineering in phenotyping. MCEs learned by using knowledge graphs outperformed MCEs learned by using EHR data in retrieving phenotype-relevant concepts. We also found that enriching a knowledge graph by adding relationships to increase connectivity of the knowledge graph improves the performance of MCEs in retrieving phenotype-relevant concepts. Future works include evaluating the performance of MCEs using the concepts from several different domains and many other embedding methods.

## Data Availability

The source code is publicly available at https://github.com/WengLab-InformaticsResearch/mcephe

https://github.com/WengLab-InformaticsResearch/concept-recommender

## CONTRIBUTORS

JL and CL implemented the methods and conducted all the experiments. JHK and AB contributed to conducting experiments and evaluation of the results. NS, CP, KN, and PR contributed to generating dataset and implementing OMOP CDM for PheKB phenotypes. CT and CW co-supervised the research and edited the manuscript. All authors were involved in developing the ideas and drafting the paper.

## FUNDING

This work was supported by National Library of Medicine grants R01LM009886-11 and 1R01LM012895-03, National Human Genome Research Institute grant 2U01-HG008680-05, and National Center for Advancing Translational Science grant 1OT2TR003434-01.

## COMPETING INTERESTS

The authors have no competing interests to declare.

## SUPPLEMENTARY MATERIAL

Supplementary material is available at *Journal of the American Medical Informatics Association*

online.

## ACKNOWLEDGEMENT

This study received institutional review board approval (AAAD1873) with a waiver for informed consent. We would like to thank eMERGE phenotyping workgroup who inspired this study.

## Notes

### Competing Interest Statement

The authors have declared no competing interest.

### Author Declarations

This study received institutional review board approval with a waiver for informed consent.

