## Supplementary for "Comparative Effectiveness of Medical Concept Embedding for Feature Engineering in Phenotyping"

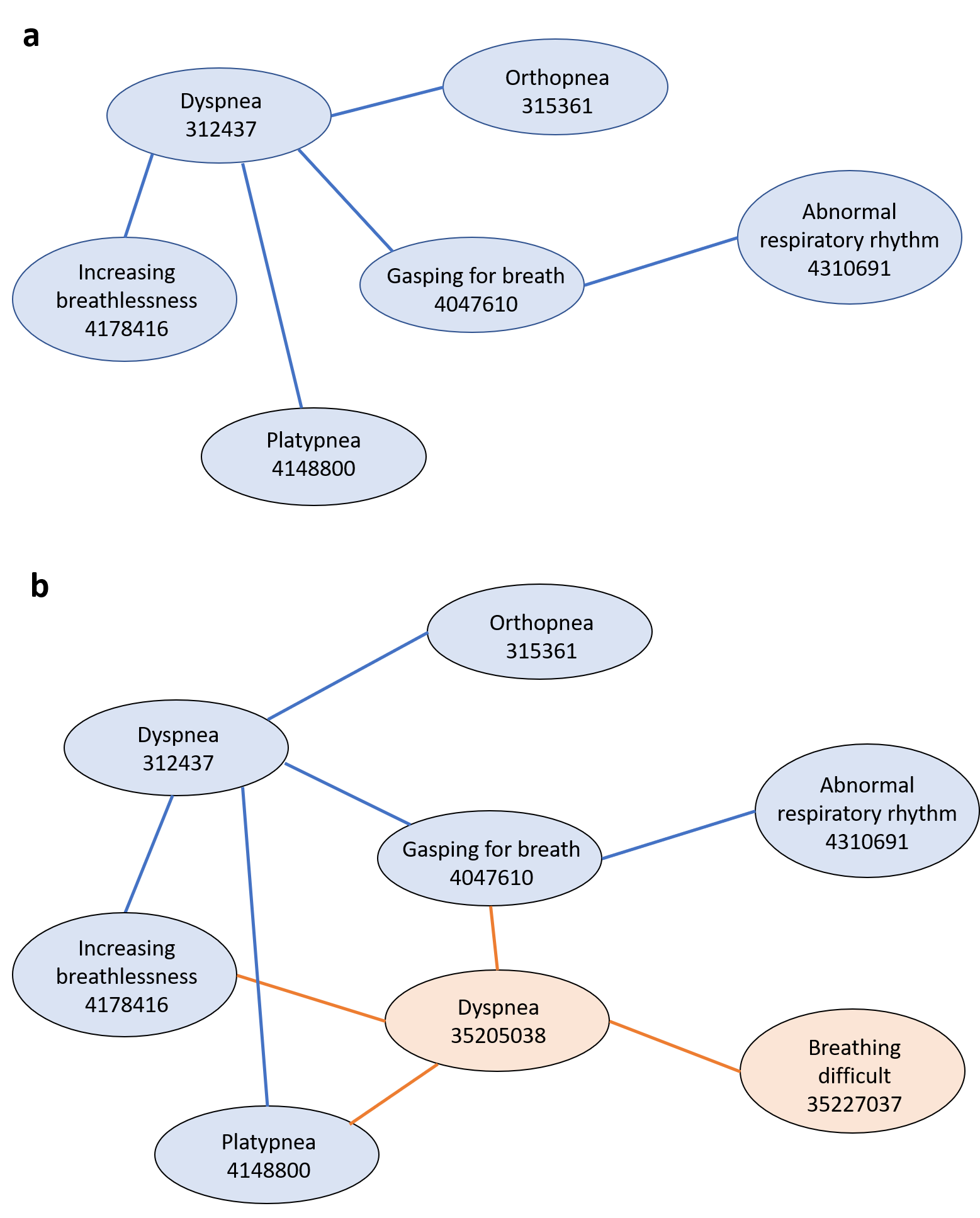

**Supplementary Figure 1.** An example of (**a**) the hierarchical knowledge graph and (**b**) the enriched knowledge graph. We depicted only a fraction of all relationships for each concept node. The number in each concept node denotes concept id in Observational Medical Outcomes Partnership common data model (OMOP CDM). Standard and non-standard concept nodes are highlighted in blue and orange respectively. “is-a” and “subsume” relationships are depicted with blue lines. Additional relationships from *concept_ancestor* table for the enriched knowledge graph are depicted with orange lines.

**Supplementary Table 1.** The number of concepts in phenotype.

| **Phenotype (abbreviation)** | **# of concepts** |
| --- | --- |
| Lipids | 637 |
| Benign Prostatic Hyperplasia (BPH) | 32 |
| Appendicitis | 50 |
| Colorectal Cancer (CRC) | 868 |
| Cardiac Conduction (QRS) | 353 |
| Autism | 417 |
| Hypothyroidism | 75 |
| Dementia | 24 |
| Red Blood Cell Indices (RBC) | 638 |
| Height | 187 |
| Chronic Kidney Disease (CKD) | 1416 |
| Resistant Hypertension (ResHTN) | 101 |
| Extreme Obesity (EO) | 1619 |
| Chronic Rhinosinusitis (CR) | 122 |
| Type 2 Diabetes Mellitus (T2DM) | 62 |
| Zoster | 253 |
| Attention Deficit Hyperactivity Disorder (ADHD) | 549 |
| Diverticulosis | 5 |
| Cardiorespiratory Fitness (CF) | 63 |
| Early Childhood Obesity (ECO) | 63 |
| Statin and MACE (Satin/MACE) | 13 |
| Epilepsy | 13 |
| HeartFailure (HF) | 107 |
| Cervical Intraepithelial Neoplasia (CIN) | 60 |
| Atopic Dermatitis (AD) | 214 |
| Diabeters/Hypertension associated Chronic Kidney Disease (DHaCKD) | 27 |
| Asthma | 150 |
| Abdominal Aortic Aneurysm (AAA) | 12 |
| ACE Inhibitor induced Cough (ACECough) | 47 |
| Venous Thromboembolism (VTE) | 784 |
| Gastroesophageal Reflux Disease (GERD) | 184 |
| Clostridium Difficile Colitis (Cdiff) | 244 |
| Community Associated Methicillin-resistant Staphylococcus Aureus (CaMRSA) | 45 |

**Supplementary Table 2.** *Hits@100%* of individual phenotypes based on a single seed concept for graph-based MCEs.

|  | n2vEmb | n2vEmb+ | LINEEmb | LINEEmb+ | SVDEmb | SVDEmb+ |
| --- | --- | --- | --- | --- | --- | --- |
| VTE | 0.339606 | 0.353887 | 0.151458 | 0.161004 | 0.151978 | 0.146946 |
| Dementia | 0.5 | 0.590278 | 0.088542 | 0.107639 | 0.003472 | 0.012153 |
| Zoster | 0.678295 | 0.725617 | 0.154181 | 0.417644 | 0.050259 | 0.049009 |
| DHaCKD | 0.22929 | 0.251479 | 0.051775 | 0.161243 | 0.013314 | 0.008876 |
| EO | 0.405445 | 0.402028 | 0.29906 | 0.318651 | 0.310645 | 0.309928 |
| CF | 0.263291 | 0.319224 | 0.073318 | 0.151172 | 0.01159 | 0.012094 |
| Height | 0.272498 | 0.279648 | 0.060768 | 0.151248 | 0.038777 | 0.036061 |
| Asthma | 0.216089 | 0.228622 | 0.042711 | 0.063733 | 0.029778 | 0.031244 |
| HF | 0.23665 | 0.264863 | 0.063368 | 0.139106 | 0.016732 | 0.020737 |
| CaMRSA | 0.167407 | 0.190123 | 0.057284 | 0.078519 | 0.010864 | 0.010864 |
| Hypothyroidism | 0.359467 | 0.352 | 0.052622 | 0.094578 | 0.011022 | 0.016711 |
| Lipids | 0.654691 | 0.675443 | 0.142975 | 0.210326 | 0.121218 | 0.125581 |
| ECO | 0.16125 | 0.184177 | 0.066264 | 0.109095 | 0.015117 | 0.018644 |
| Autism | 0.570295 | 0.604434 | 0.098107 | 0.1142 | 0.080581 | 0.077426 |
| Satin/MACE | 0.473373 | 0.597633 | 0.136095 | 0.39645 | 0 | 0.005917 |
| ADHD | 0.33235 | 0.348806 | 0.1138 | 0.143335 | 0.103677 | 0.102794 |
| RBC | 0.30977 | 0.322108 | 0.144468 | 0.208248 | 0.125102 | 0.125407 |
| AD | 0.208726 | 0.221921 | 0.058495 | 0.102572 | 0.040829 | 0.040183 |
| T2D | 0.208815 | 0.221983 | 0.064499 | 0.105348 | 0.014781 | 0.014512 |
| BPH | 0.296875 | 0.324219 | 0.052734 | 0.095703 | 0.003906 | 0.020508 |
| GERD | 0.592642 | 0.604169 | 0.067664 | 0.149482 | 0.036967 | 0.036848 |
| Appendicitis | 0.144 | 0.1632 | 0.0324 | 0.0476 | 0.0184 | 0.0104 |
| Epilepsy | 0.662722 | 0.692308 | 0.118343 | 0.088757 | 0 | 0.047337 |
| CRC | 0.691191 | 0.726809 | 0.170265 | 0.218444 | 0.168087 | 0.169422 |
| CIN | 0.213732 | 0.244757 | 0.074404 | 0.147371 | 0.012065 | 0.011491 |
| CR | 0.39765 | 0.442661 | 0.058261 | 0.088109 | 0.02172 | 0.024725 |
| CKD | 0.615831 | 0.63797 | 0.343286 | 0.369866 | 0.271428 | 0.267425 |
| ResHTN | 0.140869 | 0.180374 | 0.051368 | 0.10097 | 0.016861 | 0.023625 |
| Cdiff | 0.37505 | 0.378544 | 0.064969 | 0.102593 | 0.048945 | 0.048878 |
| QRS | 0.315833 | 0.320119 | 0.077641 | 0.098673 | 0.067302 | 0.069739 |
| ACECough | 0.37483 | 0.389316 | 0.086012 | 0.128565 | 0.012223 | 0.013128 |
| Diverticulosis | 0.28 | 0.2 | 0.08 | 0.08 | 0 | 0 |
| AAA | 0.819444 | 0.902778 | 0.25 | 0.229167 | 0.006944 | 0.020833 |

**Supplementary Table 3.** *Hits@100%* of individual phenotypes based on a single seed concept for EHR-based MCEs

|  | GloVeEmb_V | GloVeEmb_5Y | SGEmb_V | SGEmb_5Y |
| --- | --- | --- | --- | --- |
| VTE | 0.309767 | 0.380908 | 0.198834 | 0.162009 |
| Dementia | 0.05482 | 0.151229 | 0.035917 | 0 |
| Zoster | 0.17601 | 0.15219 | 0.072483 | 0.046344 |
| DHaCKD | 0.14645 | 0.223373 | 0.097633 | 0.065089 |
| EO | 0.348938 | 0.361097 | 0.305231 | 0.345157 |
| CF | 0.250278 | 0.387222 | 0.206944 | 0.095833 |
| Height | 0.080867 | 0.073622 | 0.055408 | 0.038112 |
| Asthma | 0.109471 | 0.130835 | 0.10354 | 0.052083 |
| HF | 0.227973 | 0.312717 | 0.167969 | 0.097873 |
| CaMRSA | 0.113952 | 0.119065 | 0.042367 | 0.019722 |
| Hypothyroidism | 0.090332 | 0.134033 | 0.046387 | 0.019531 |
| Lipids | 0.29871 | 0.209319 | 0.157895 | 0.116879 |
| ECO | 0.053212 | 0.07928 | 0.027143 | 0.016931 |
| Autism | 0.329754 | 0.365215 | 0.157369 | 0.098104 |
| Satin/MACE | 0.097222 | 0.222222 | 0.090278 | 0 |
| ADHD | 0.152854 | 0.207358 | 0.143678 | 0.116851 |
| RBC | 0.234899 | 0.193579 | 0.184776 | 0.13219 |
| AD | 0.173956 | 0.18004 | 0.099028 | 0.054921 |
| T2D | 0.08341 | 0.126193 | 0.062173 | 0.020006 |
| BPH | 0.128472 | 0.173611 | 0.029514 | 0.005208 |
| GERD | 0.101094 | 0.099379 | 0.063242 | 0.043247 |
| Appendicitis | 0.057039 | 0.073336 | 0.055229 | 0.020371 |
| Epilepsy | 0.378698 | 0.414201 | 0.047337 | 0.005917 |
| CRC | 0.3167 | 0.251813 | 0.215919 | 0.173736 |
| CIN | 0.2283 | 0.257134 | 0.143579 | 0.068668 |
| CR | 0.178612 | 0.220124 | 0.096596 | 0.047903 |
| CKD | 0.179311 | 0.209123 | 0.115733 | 0.103321 |
| ResHTN | 0.085877 | 0.094464 | 0.054433 | 0.031444 |
| Cdiff | 0.219423 | 0.199882 | 0.081941 | 0.0444 |
| QRS | 0.314882 | 0.340027 | 0.243802 | 0.179345 |
| ACECough | 0.105388 | 0.137996 | 0.056711 | 0.031664 |
| Diverticulosis | 0.111111 | 0 | 0 | 0 |
| AAA | 0.201389 | 0.395833 | 0.090278 | 0 |

**Supplementary Table 4.** *Hits@200%* of individual phenotypes based on a single seed concept for graph-based MCEs

|  | n2vEmb | n2vEmb+ | LINEEmb | LINEEmb+ | SVDEmb | SVDEmb+ |
| --- | --- | --- | --- | --- | --- | --- |
| VTE | 0.490485 | 0.508096 | 0.29704 | 0.304594 | 0.303825 | 0.299692 |
| Dementia | 0.711806 | 0.746528 | 0.098958 | 0.125 | 0.003472 | 0.015625 |
| Zoster | 0.820572 | 0.835851 | 0.229358 | 0.52527 | 0.100908 | 0.099111 |
| DHaCKD | 0.328402 | 0.397929 | 0.072485 | 0.199704 | 0.016272 | 0.014793 |
| EO | 0.656327 | 0.652706 | 0.605487 | 0.618988 | 0.621433 | 0.621536 |
| CF | 0.386747 | 0.410935 | 0.089443 | 0.177375 | 0.023936 | 0.024439 |
| Height | 0.321056 | 0.342704 | 0.097429 | 0.193486 | 0.074924 | 0.070663 |
| Asthma | 0.331111 | 0.344711 | 0.074089 | 0.098578 | 0.059156 | 0.059733 |
| HF | 0.347544 | 0.351816 | 0.090958 | 0.172036 | 0.03471 | 0.045034 |
| CaMRSA | 0.238025 | 0.272099 | 0.07358 | 0.099259 | 0.015309 | 0.020741 |
| Hypothyroidism | 0.4 | 0.402489 | 0.069867 | 0.116978 | 0.0224 | 0.027556 |
| Lipids | 0.802725 | 0.823014 | 0.260559 | 0.329516 | 0.241748 | 0.253402 |
| ECO | 0.221466 | 0.251701 | 0.097254 | 0.147392 | 0.024691 | 0.028723 |
| Autism | 0.71756 | 0.775165 | 0.180202 | 0.198872 | 0.157926 | 0.153881 |
| Satin/MACE | 0.538462 | 0.739645 | 0.16568 | 0.43787 | 0.011834 | 0.005917 |
| ADHD | 0.472763 | 0.510329 | 0.215408 | 0.246273 | 0.206635 | 0.205799 |
| RBC | 0.418736 | 0.44138 | 0.266305 | 0.334146 | 0.248246 | 0.250204 |
| AD | 0.270959 | 0.281083 | 0.100948 | 0.148029 | 0.082169 | 0.078965 |
| T2D | 0.271701 | 0.273851 | 0.088417 | 0.133566 | 0.023918 | 0.024187 |
| BPH | 0.376953 | 0.435547 | 0.074219 | 0.120117 | 0.010742 | 0.033203 |
| GERD | 0.668548 | 0.676013 | 0.104183 | 0.203948 | 0.071337 | 0.071546 |
| Appendicitis | 0.1872 | 0.208 | 0.046 | 0.064 | 0.0268 | 0.02 |
| Epilepsy | 0.881657 | 0.899408 | 0.136095 | 0.106509 | 0.005917 | 0.047337 |
| CRC | 0.847108 | 0.871316 | 0.328363 | 0.371998 | 0.334379 | 0.337532 |
| CIN | 0.265154 | 0.30135 | 0.091066 | 0.170353 | 0.019247 | 0.021833 |
| CR | 0.524418 | 0.583567 | 0.087153 | 0.122055 | 0.045625 | 0.047879 |
| CKD | 0.819447 | 0.828238 | 0.621276 | 0.642648 | 0.543775 | 0.540574 |
| ResHTN | 0.188217 | 0.233997 | 0.07362 | 0.124007 | 0.034114 | 0.045192 |
| Cdiff | 0.647071 | 0.658963 | 0.109682 | 0.15436 | 0.096967 | 0.096799 |
| QRS | 0.432036 | 0.438364 | 0.140907 | 0.161561 | 0.132885 | 0.141424 |
| ACECough | 0.564962 | 0.59665 | 0.103214 | 0.145767 | 0.020371 | 0.020824 |
| Diverticulosis | 0.4 | 0.36 | 0.08 | 0.08 | 0 | 0 |
| AAA | 0.895833 | 0.916667 | 0.270833 | 0.270833 | 0.006944 | 0.020833 |

**Supplementary Table 5.** *Hits@200%* of individual phenotypes based on a single seed concept for EHR-based MCEs

|  | GloVeEmb_V | GloVeEmb_5Y | SGEmb_V | SGEmb_5Y |
| --- | --- | --- | --- | --- |
| VTE | 0.418458 | 0.496456 | 0.309952 | 0.333992 |
| Dementia | 0.081285 | 0.217391 | 0.081285 | 0.009452 |
| Zoster | 0.265861 | 0.20077 | 0.123759 | 0.092774 |
| DHaCKD | 0.220414 | 0.33284 | 0.155325 | 0.102071 |
| EO | 0.665879 | 0.674046 | 0.654565 | 0.700953 |
| CF | 0.402222 | 0.558056 | 0.300833 | 0.179444 |
| Height | 0.129541 | 0.124439 | 0.102602 | 0.073316 |
| Asthma | 0.198302 | 0.228636 | 0.184076 | 0.102382 |
| HF | 0.378798 | 0.479818 | 0.25803 | 0.186089 |
| CaMRSA | 0.145362 | 0.157049 | 0.067202 | 0.029949 |
| Hypothyroidism | 0.129639 | 0.185303 | 0.078125 | 0.044678 |
| Lipids | 0.449511 | 0.331709 | 0.292146 | 0.239487 |
| ECO | 0.077399 | 0.116367 | 0.048105 | 0.031443 |
| Autism | 0.435998 | 0.487837 | 0.258885 | 0.201841 |
| Satin/MACE | 0.131944 | 0.291667 | 0.131944 | 0.006944 |
| ADHD | 0.276305 | 0.352579 | 0.27894 | 0.24385 |
| RBC | 0.402013 | 0.344685 | 0.326159 | 0.268372 |
| AD | 0.253481 | 0.26844 | 0.165453 | 0.112419 |
| T2D | 0.138504 | 0.191444 | 0.091105 | 0.044014 |
| BPH | 0.164931 | 0.241319 | 0.045139 | 0.013889 |
| GERD | 0.15 | 0.161221 | 0.110703 | 0.087574 |
| Appendicitis | 0.089181 | 0.109099 | 0.093708 | 0.036215 |
| Epilepsy | 0.449704 | 0.585799 | 0.08284 | 0.005917 |
| CRC | 0.515552 | 0.449677 | 0.39254 | 0.358764 |
| CIN | 0.349287 | 0.390904 | 0.217004 | 0.143876 |
| CR | 0.253016 | 0.316791 | 0.151393 | 0.10507 |
| CKD | 0.3093 | 0.335125 | 0.215451 | 0.196537 |
| ResHTN | 0.130268 | 0.141895 | 0.093539 | 0.065398 |
| Cdiff | 0.340892 | 0.29582 | 0.135718 | 0.092775 |
| QRS | 0.500014 | 0.513706 | 0.370506 | 0.30192 |
| ACECough | 0.163989 | 0.194707 | 0.079395 | 0.059546 |
| Diverticulosis | 0.111111 | 0.111111 | 0 | 0 |
| AAA | 0.243056 | 0.458333 | 0.104167 | 0 |

**Supplementary Table 6.** *Hits@500%* of individual phenotypes based on a single seed concept for graph-based MCEs

|  | n2vEmb | n2vEmb+ | LINEEmb | LINEEmb+ | SVDEmb | SVDEmb+ |
| --- | --- | --- | --- | --- | --- | --- |
| VTE | 0.814999 | 0.822943 | 0.743098 | 0.741807 | 0.757596 | 0.750485 |
| Dementia | 0.871528 | 0.871528 | 0.126736 | 0.164931 | 0.013889 | 0.026042 |
| Zoster | 0.875924 | 0.874799 | 0.401084 | 0.688372 | 0.250808 | 0.252574 |
| DHaCKD | 0.445266 | 0.45858 | 0.10355 | 0.25 | 0.039941 | 0.031065 |
| EO | 0.999382 | 0.999382 | 0.999382 | 0.999382 | 0.999422 | 0.99943 |
| CF | 0.564878 | 0.557319 | 0.141597 | 0.235576 | 0.066264 | 0.067019 |
| Height | 0.41654 | 0.453087 | 0.208842 | 0.297035 | 0.180589 | 0.175899 |
| Asthma | 0.508489 | 0.525022 | 0.157111 | 0.183067 | 0.146933 | 0.149467 |
| HF | 0.513528 | 0.508188 | 0.160021 | 0.246796 | 0.094963 | 0.112051 |
| CaMRSA | 0.354074 | 0.380741 | 0.119012 | 0.141235 | 0.040494 | 0.055309 |
| Hypothyroidism | 0.437689 | 0.456533 | 0.118044 | 0.172978 | 0.0592 | 0.068267 |
| Lipids | 0.878382 | 0.892123 | 0.606849 | 0.645831 | 0.610832 | 0.625017 |
| ECO | 0.320988 | 0.346939 | 0.156715 | 0.223986 | 0.05165 | 0.069287 |
| Autism | 0.868054 | 0.917056 | 0.413271 | 0.436148 | 0.392006 | 0.388319 |
| Satin/MACE | 0.615385 | 0.781065 | 0.254438 | 0.502959 | 0.023669 | 0.011834 |
| ADHD | 0.73565 | 0.78652 | 0.517648 | 0.544148 | 0.519834 | 0.521468 |
| RBC | 0.696053 | 0.72572 | 0.622235 | 0.666424 | 0.616572 | 0.6214 |
| AD | 0.419322 | 0.422526 | 0.217916 | 0.265619 | 0.203253 | 0.193619 |
| T2D | 0.340769 | 0.348025 | 0.13061 | 0.182209 | 0.056436 | 0.061274 |
| BPH | 0.513672 | 0.634766 | 0.105469 | 0.161133 | 0.027344 | 0.054688 |
| GERD | 0.732957 | 0.740781 | 0.213294 | 0.333931 | 0.175013 | 0.177521 |
| Appendicitis | 0.294 | 0.3068 | 0.0804 | 0.1056 | 0.0568 | 0.0528 |
| Epilepsy | 0.905325 | 0.923077 | 0.159763 | 0.153846 | 0.011834 | 0.047337 |
| CRC | 0.963406 | 0.966954 | 0.818318 | 0.827228 | 0.835215 | 0.835381 |
| CIN | 0.335248 | 0.382074 | 0.127837 | 0.218328 | 0.048549 | 0.061477 |
| CR | 0.676798 | 0.738611 | 0.161464 | 0.207841 | 0.110648 | 0.120005 |
| CKD | 0.999291 | 0.999291 | 0.999291 | 0.999291 | 0.999332 | 0.999355 |
| ResHTN | 0.282031 | 0.320067 | 0.128517 | 0.180571 | 0.082639 | 0.105284 |
| Cdiff | 0.925692 | 0.951172 | 0.235538 | 0.284265 | 0.235891 | 0.2371 |
| QRS | 0.637034 | 0.633345 | 0.330215 | 0.346058 | 0.335389 | 0.359625 |
| ACECough | 0.756904 | 0.780444 | 0.140788 | 0.183794 | 0.049796 | 0.043006 |
| Diverticulosis | 0.6 | 0.52 | 0.12 | 0.08 | 0 | 0 |
| AAA | 0.916667 | 0.916667 | 0.347222 | 0.3125 | 0.013889 | 0.020833 |

**Supplementary Table 7.** *Hits@500%* of individual phenotypes based on a single seed concept for EHR-based MCEs

|  | GloVeEmb_V | GloVeEmb_5Y | SGEmb_V | SGEmb_5Y |
| --- | --- | --- | --- | --- |
| VTE | 0.885081 | 0.9046 | 0.847901 | 0.877323 |
| Dementia | 0.166352 | 0.294896 | 0.160681 | 0.020794 |
| Zoster | 0.420487 | 0.314113 | 0.247046 | 0.233109 |
| DHaCKD | 0.359467 | 0.508876 | 0.263314 | 0.18787 |
| EO | 0.999219 | 0.999227 | 0.999219 | 0.999227 |
| CF | 0.663889 | 0.745 | 0.441111 | 0.322222 |
| Height | 0.254235 | 0.254898 | 0.215867 | 0.183469 |
| Asthma | 0.436391 | 0.442853 | 0.358893 | 0.242622 |
| HF | 0.638889 | 0.744792 | 0.401476 | 0.344727 |
| CaMRSA | 0.20599 | 0.234478 | 0.124909 | 0.077429 |
| Hypothyroidism | 0.203613 | 0.304688 | 0.166748 | 0.110107 |
| Lipids | 0.759991 | 0.714522 | 0.637764 | 0.629858 |
| ECO | 0.148885 | 0.21204 | 0.099167 | 0.077667 |
| Autism | 0.692667 | 0.729733 | 0.565698 | 0.517105 |
| Satin/MACE | 0.229167 | 0.479167 | 0.1875 | 0.013889 |
| ADHD | 0.680721 | 0.726765 | 0.644239 | 0.644643 |
| RBC | 0.749449 | 0.749322 | 0.691538 | 0.694207 |
| AD | 0.435912 | 0.474838 | 0.330853 | 0.275614 |
| T2D | 0.243767 | 0.305017 | 0.15574 | 0.122191 |
| BPH | 0.217014 | 0.303819 | 0.067708 | 0.039931 |
| GERD | 0.277266 | 0.304348 | 0.234219 | 0.210486 |
| Appendicitis | 0.179719 | 0.204617 | 0.168855 | 0.093708 |
| Epilepsy | 0.514793 | 0.686391 | 0.142012 | 0.017751 |
| CRC | 0.958489 | 0.969101 | 0.946516 | 0.939993 |
| CIN | 0.534185 | 0.598989 | 0.336207 | 0.373068 |
| CR | 0.401537 | 0.48578 | 0.274275 | 0.22946 |
| CKD | 0.598687 | 0.636388 | 0.484508 | 0.488499 |
| ResHTN | 0.272163 | 0.25512 | 0.192231 | 0.165147 |
| Cdiff | 0.506509 | 0.461465 | 0.283419 | 0.234708 |
| QRS | 0.79337 | 0.822475 | 0.729558 | 0.600362 |
| ACECough | 0.275047 | 0.336484 | 0.145558 | 0.15879 |
| Diverticulosis | 0.111111 | 0.111111 | 0 | 0 |
| AAA | 0.277778 | 0.555556 | 0.152778 | 0 |

**Supplementary Table 8.** *Hits@1000%* of individual phenotypes based on a single seed concept for graph-based MCEs

|  | n2vEmb | n2vEmb+ | LINEEmb | LINEEmb+ | SVDEmb | SVDEmb+ |
| --- | --- | --- | --- | --- | --- | --- |
| VTE | 0.998723 | 0.998723 | 0.998723 | 0.998723 | 0.998837 | 0.998848 |
| Dementia | 0.927083 | 0.913194 | 0.168403 | 0.210069 | 0.034722 | 0.045139 |
| Zoster | 0.917449 | 0.921948 | 0.635301 | 0.842803 | 0.491228 | 0.498258 |
| DHaCKD | 0.529586 | 0.52071 | 0.143491 | 0.310651 | 0.065089 | 0.047337 |
| EO | 0.999382 | 0.999382 | 0.999382 | 0.999382 | 0.999422 | 0.99943 |
| CF | 0.671958 | 0.681532 | 0.205593 | 0.306878 | 0.129504 | 0.125472 |
| Height | 0.532186 | 0.590752 | 0.383111 | 0.457691 | 0.357116 | 0.366553 |
| Asthma | 0.670222 | 0.672533 | 0.293733 | 0.320533 | 0.287111 | 0.283911 |
| HF | 0.650676 | 0.634834 | 0.259879 | 0.348612 | 0.194731 | 0.233624 |
| CaMRSA | 0.466173 | 0.48642 | 0.182222 | 0.194074 | 0.094321 | 0.112099 |
| Hypothyroidism | 0.467378 | 0.510222 | 0.187022 | 0.244978 | 0.126578 | 0.133156 |
| Lipids | 0.998428 | 0.998428 | 0.998428 | 0.998428 | 0.998571 | 0.998576 |
| ECO | 0.420509 | 0.424792 | 0.230285 | 0.306122 | 0.109095 | 0.126228 |
| Autism | 0.97209 | 0.98608 | 0.797765 | 0.815337 | 0.795881 | 0.78708 |
| Satin/MACE | 0.757396 | 0.781065 | 0.307692 | 0.556213 | 0.035503 | 0.029586 |
| ADHD | 0.998172 | 0.998172 | 0.998172 | 0.998172 | 0.998289 | 0.998326 |
| RBC | 0.998433 | 0.998433 | 0.998433 | 0.998433 | 0.99856 | 0.99858 |
| AD | 0.605064 | 0.615477 | 0.408041 | 0.447646 | 0.401055 | 0.385457 |
| T2D | 0.39828 | 0.416286 | 0.187046 | 0.250202 | 0.103198 | 0.114754 |
| BPH | 0.613281 | 0.786133 | 0.138672 | 0.195313 | 0.0625 | 0.084961 |
| GERD | 0.795246 | 0.804264 | 0.387142 | 0.510406 | 0.350324 | 0.357371 |
| Appendicitis | 0.4008 | 0.416 | 0.1312 | 0.1532 | 0.11 | 0.0988 |
| Epilepsy | 0.91716 | 0.923077 | 0.189349 | 0.218935 | 0.029586 | 0.053254 |
| CRC | 0.998847 | 0.998847 | 0.998847 | 0.998847 | 0.998932 | 0.998954 |
| CIN | 0.410227 | 0.450158 | 0.175524 | 0.283252 | 0.09796 | 0.113473 |
| CR | 0.794959 | 0.845502 | 0.281128 | 0.332149 | 0.233181 | 0.228946 |
| CKD | 0.999291 | 0.999291 | 0.999291 | 0.999291 | 0.999332 | 0.999355 |
| ResHTN | 0.424272 | 0.463974 | 0.226252 | 0.270758 | 0.173022 | 0.20547 |
| Cdiff | 0.975847 | 0.983724 | 0.448821 | 0.481389 | 0.467566 | 0.476871 |
| QRS | 0.867478 | 0.856082 | 0.65508 | 0.661972 | 0.673634 | 0.691406 |
| ACECough | 0.81847 | 0.807605 | 0.189226 | 0.227252 | 0.090086 | 0.094613 |
| Diverticulosis | 0.64 | 0.68 | 0.16 | 0.08 | 0 | 0 |
| AAA | 0.916667 | 0.916667 | 0.409722 | 0.375 | 0.034722 | 0.027778 |

**Supplementary Table 9.** *Hits@1000%* of individual phenotypes based on a single seed concept for EHR-based MCEs

|  | GloVeEmb_V | GloVeEmb_5Y | SGEmb_V | SGEmb_5Y |
| --- | --- | --- | --- | --- |
| VTE | 0.998433 | 0.998452 | 0.998433 | 0.998452 |
| Dementia | 0.31758 | 0.385633 | 0.232514 | 0.062382 |
| Zoster | 0.612329 | 0.506479 | 0.430529 | 0.48724 |
| DHaCKD | 0.575444 | 0.670118 | 0.381657 | 0.323964 |
| EO | 0.999219 | 0.999227 | 0.999219 | 0.999227 |
| CF | 0.816667 | 0.895556 | 0.567222 | 0.471389 |
| Height | 0.433929 | 0.450561 | 0.423316 | 0.363265 |
| Asthma | 0.669416 | 0.678675 | 0.608796 | 0.44565 |
| HF | 0.794705 | 0.904622 | 0.544379 | 0.52474 |
| CaMRSA | 0.28634 | 0.357925 | 0.223521 | 0.145362 |
| Hypothyroidism | 0.286865 | 0.460693 | 0.250488 | 0.215332 |
| Lipids | 0.997872 | 0.99789 | 0.997872 | 0.99789 |
| ECO | 0.241602 | 0.342112 | 0.184897 | 0.163128 |
| Autism | 0.997416 | 0.997423 | 0.997416 | 0.997423 |
| Satin/MACE | 0.354167 | 0.604167 | 0.256944 | 0.048611 |
| ADHD | 0.997899 | 0.997904 | 0.997899 | 0.997904 |
| RBC | 0.998043 | 0.998058 | 0.998043 | 0.998058 |
| AD | 0.681395 | 0.701695 | 0.619938 | 0.554682 |
| T2D | 0.329024 | 0.405355 | 0.219144 | 0.237919 |
| BPH | 0.258681 | 0.355903 | 0.109375 | 0.076389 |
| GERD | 0.504961 | 0.498476 | 0.44918 | 0.424829 |
| Appendicitis | 0.29244 | 0.334541 | 0.274785 | 0.203712 |
| Epilepsy | 0.591716 | 0.763314 | 0.218935 | 0.035503 |
| CRC | 0.998538 | 0.998547 | 0.998538 | 0.998547 |
| CIN | 0.672711 | 0.767539 | 0.456599 | 0.504756 |
| CR | 0.578426 | 0.675093 | 0.467251 | 0.417983 |
| CKD | 0.982981 | 0.982795 | 0.985171 | 0.9826 |
| ResHTN | 0.487383 | 0.421984 | 0.341525 | 0.300436 |
| Cdiff | 0.698917 | 0.695165 | 0.51152 | 0.491084 |
| QRS | 0.982289 | 0.981014 | 0.979448 | 0.947363 |
| ACECough | 0.420605 | 0.485822 | 0.239603 | 0.227316 |
| Diverticulosis | 0.111111 | 0.111111 | 0 | 0 |
| AAA | 0.354167 | 0.666667 | 0.1875 | 0.013889 |

**Supplementary Table 10.** *Hits@2000%* of individual phenotypes based on a single seed concept for graph-based MCEs

|  | n2vEmb | n2vEmb+ | LINEEmb | LINEEmb+ | SVDEmb | SVDEmb+ |
| --- | --- | --- | --- | --- | --- | --- |
| VTE | 0.998723 | 0.998723 | 0.998723 | 0.998723 | 0.998837 | 0.998848 |
| Dementia | 0.942708 | 0.951389 | 0.225694 | 0.276042 | 0.076389 | 0.086806 |
| Zoster | 0.994923 | 0.993079 | 0.982174 | 0.99222 | 0.970645 | 0.970473 |
| DHaCKD | 0.62426 | 0.597633 | 0.214497 | 0.386095 | 0.091716 | 0.100592 |
| EO | 0.999382 | 0.999382 | 0.999382 | 0.999382 | 0.999422 | 0.99943 |
| CF | 0.774502 | 0.801713 | 0.309146 | 0.411691 | 0.251197 | 0.243638 |
| Height | 0.762561 | 0.823472 | 0.729904 | 0.759787 | 0.71143 | 0.717493 |
| Asthma | 0.858089 | 0.844667 | 0.563778 | 0.5852 | 0.572089 | 0.551067 |
| HF | 0.81675 | 0.797526 | 0.457102 | 0.521093 | 0.388305 | 0.435564 |
| CaMRSA | 0.604938 | 0.584198 | 0.291358 | 0.285926 | 0.173827 | 0.215309 |
| Hypothyroidism | 0.516978 | 0.596622 | 0.323556 | 0.375111 | 0.283378 | 0.289244 |
| Lipids | 0.998428 | 0.998428 | 0.998428 | 0.998428 | 0.998571 | 0.998576 |
| ECO | 0.555808 | 0.526833 | 0.351474 | 0.433862 | 0.207861 | 0.233812 |
| Autism | 0.997596 | 0.997596 | 0.997596 | 0.997596 | 0.997781 | 0.99785 |
| Satin/MACE | 0.905325 | 0.792899 | 0.390533 | 0.60355 | 0.065089 | 0.071006 |
| ADHD | 0.998172 | 0.998172 | 0.998172 | 0.998172 | 0.998289 | 0.998326 |
| RBC | 0.998433 | 0.998433 | 0.998433 | 0.998433 | 0.99856 | 0.99858 |
| AD | 0.881052 | 0.892577 | 0.797726 | 0.813301 | 0.802376 | 0.812589 |
| T2D | 0.514647 | 0.542596 | 0.298844 | 0.356625 | 0.215533 | 0.241333 |
| BPH | 0.750977 | 0.920898 | 0.19043 | 0.251953 | 0.126953 | 0.135742 |
| GERD | 0.911523 | 0.903998 | 0.72582 | 0.802144 | 0.688047 | 0.686852 |
| Appendicitis | 0.51 | 0.5176 | 0.23 | 0.2472 | 0.2056 | 0.1892 |
| Epilepsy | 0.91716 | 0.923077 | 0.254438 | 0.284024 | 0.047337 | 0.059172 |
| CRC | 0.998847 | 0.998847 | 0.998847 | 0.998847 | 0.998932 | 0.998954 |
| CIN | 0.513933 | 0.51422 | 0.264866 | 0.38667 | 0.193335 | 0.270899 |
| CR | 0.897343 | 0.9429 | 0.508572 | 0.550031 | 0.46561 | 0.444778 |
| CKD | 0.999291 | 0.999291 | 0.999291 | 0.999291 | 0.999332 | 0.999355 |
| ResHTN | 0.664739 | 0.68111 | 0.403196 | 0.429076 | 0.367219 | 0.407607 |
| Cdiff | 0.99523 | 0.995583 | 0.916454 | 0.914069 | 0.933032 | 0.934342 |
| QRS | 0.997159 | 0.997159 | 0.997159 | 0.997159 | 0.997433 | 0.997442 |
| ACECough | 0.883658 | 0.845632 | 0.279765 | 0.299683 | 0.173834 | 0.178814 |
| Diverticulosis | 0.68 | 0.76 | 0.2 | 0.08 | 0 | 0 |
| AAA | 0.916667 | 0.916667 | 0.458333 | 0.409722 | 0.048611 | 0.034722 |

**Supplementary Table 11.** *Hits@2000%* of individual phenotypes based on a single seed concept for EHR-based MCEs

|  | GloVeEmb_V | GloVeEmb_5Y | SGEmb_V | SGEmb_5Y |
| --- | --- | --- | --- | --- |
| VTE | 0.998433 | 0.998452 | 0.998433 | 0.998452 |
| Dementia | 0.432892 | 0.533081 | 0.311909 | 0.128544 |
| Zoster | 0.994565 | 0.994681 | 0.994565 | 0.994681 |
| DHaCKD | 0.764793 | 0.821006 | 0.507396 | 0.5 |
| EO | 0.999219 | 0.999227 | 0.999219 | 0.999227 |
| CF | 0.931667 | 0.972222 | 0.728889 | 0.638056 |
| Height | 0.72801 | 0.786429 | 0.738724 | 0.742449 |
| Asthma | 0.924672 | 0.945264 | 0.904225 | 0.822145 |
| HF | 0.945964 | 0.977322 | 0.841254 | 0.716363 |
| CaMRSA | 0.446311 | 0.544923 | 0.391527 | 0.318481 |
| Hypothyroidism | 0.408691 | 0.63501 | 0.385254 | 0.394287 |
| Lipids | 0.997872 | 0.99789 | 0.997872 | 0.99789 |
| ECO | 0.442623 | 0.526203 | 0.367912 | 0.335931 |
| Autism | 0.997416 | 0.997423 | 0.997416 | 0.997423 |
| Satin/MACE | 0.479167 | 0.763889 | 0.375 | 0.145833 |
| ADHD | 0.997899 | 0.997904 | 0.997899 | 0.997904 |
| RBC | 0.998043 | 0.998058 | 0.998043 | 0.998058 |
| AD | 0.994845 | 0.994845 | 0.994845 | 0.994845 |
| T2D | 0.472145 | 0.504155 | 0.342875 | 0.434287 |
| BPH | 0.347222 | 0.418403 | 0.196181 | 0.118056 |
| GERD | 0.847852 | 0.897033 | 0.854766 | 0.874851 |
| Appendicitis | 0.465369 | 0.559077 | 0.47895 | 0.383431 |
| Epilepsy | 0.674556 | 0.804734 | 0.301775 | 0.08284 |
| CRC | 0.998538 | 0.998547 | 0.998538 | 0.998547 |
| CIN | 0.801427 | 0.865636 | 0.627527 | 0.666766 |
| CR | 0.809035 | 0.860529 | 0.786986 | 0.709135 |
| CKD | 0.997199 | 0.997207 | 0.997199 | 0.997207 |
| ResHTN | 0.752543 | 0.738142 | 0.665213 | 0.557141 |
| Cdiff | 0.994709 | 0.994709 | 0.994709 | 0.994709 |
| QRS | 0.996997 | 0.997006 | 0.996997 | 0.997006 |
| ACECough | 0.561437 | 0.650284 | 0.413516 | 0.428639 |
| Diverticulosis | 0.111111 | 0.111111 | 0 | 0 |
| AAA | 0.486111 | 0.763889 | 0.284722 | 0.055556 |

**Supplementary Table 12.** *Hits@100%* of individual phenotypes based on five seed concepts for graph-based MCEs

|  | n2vEmb | n2vEmb+ | LINEEmb | LINEEmb+ | SVDEmb | SVDEmb+ |
| --- | --- | --- | --- | --- | --- | --- |
| VTE | 0.467283 | 0.491821 | 0.153872 | 0.174441 | 0.152758 | 0.147851 |
| Dementia | 0.732639 | 0.798611 | 0.25 | 0.364583 | 0.048611 | 0.065972 |
| Zoster | 0.840351 | 0.862207 | 0.256761 | 0.628318 | 0.052477 | 0.05118 |
| DHaCKD | 0.276627 | 0.318047 | 0.242604 | 0.406805 | 0.08284 | 0.047337 |
| EO | 0.510689 | 0.516856 | 0.294687 | 0.336514 | 0.31126 | 0.311584 |
| CF | 0.451499 | 0.555556 | 0.15999 | 0.31998 | 0.02973 | 0.031242 |
| Height | 0.470302 | 0.496154 | 0.092854 | 0.275873 | 0.042152 | 0.043324 |
| Asthma | 0.398178 | 0.422222 | 0.079956 | 0.106444 | 0.038711 | 0.037911 |
| HF | 0.334728 | 0.374422 | 0.125845 | 0.26246 | 0.034176 | 0.027946 |
| CaMRSA | 0.362963 | 0.352593 | 0.178765 | 0.18963 | 0.045926 | 0.028642 |
| Hypothyroidism | 0.518044 | 0.5504 | 0.122667 | 0.193956 | 0.023644 | 0.029156 |
| Lipids | 0.809091 | 0.823773 | 0.175221 | 0.310332 | 0.121658 | 0.129826 |
| ECO | 0.302091 | 0.351222 | 0.167045 | 0.223986 | 0.036785 | 0.04006 |
| Autism | 0.782203 | 0.820908 | 0.114298 | 0.150685 | 0.080673 | 0.08295 |
| Satin/MACE | 0.751479 | 0.810651 | 0.52071 | 0.721893 | 0.076923 | 0.094675 |
| ADHD | 0.53967 | 0.566604 | 0.122476 | 0.1909 | 0.1061 | 0.107316 |
| RBC | 0.462373 | 0.474133 | 0.179133 | 0.29417 | 0.125072 | 0.127333 |
| AD | 0.389396 | 0.411846 | 0.088443 | 0.168944 | 0.045879 | 0.042876 |
| T2D | 0.37678 | 0.389411 | 0.159366 | 0.215265 | 0.038699 | 0.026874 |
| BPH | 0.513672 | 0.591797 | 0.172852 | 0.238281 | 0.041016 | 0.046875 |
| GERD | 0.786407 | 0.776643 | 0.101138 | 0.265012 | 0.042014 | 0.045627 |
| Appendicitis | 0.2848 | 0.3692 | 0.1224 | 0.1372 | 0.034 | 0.0316 |
| Epilepsy | 0.751479 | 0.757396 | 0.408284 | 0.550296 | 0.076923 | 0.106509 |
| CRC | 0.881253 | 0.902587 | 0.183126 | 0.272564 | 0.168801 | 0.17305 |
| CIN | 0.267739 | 0.259408 | 0.152542 | 0.322321 | 0.035909 | 0.026716 |
| CR | 0.619562 | 0.678096 | 0.104296 | 0.16406 | 0.031145 | 0.03661 |
| CKD | 0.756799 | 0.777695 | 0.404663 | 0.440422 | 0.272621 | 0.266344 |
| ResHTN | 0.279384 | 0.335065 | 0.102245 | 0.170572 | 0.029997 | 0.027056 |
| Cdiff | 0.467532 | 0.475107 | 0.087896 | 0.164086 | 0.054152 | 0.053463 |
| QRS | 0.476554 | 0.489347 | 0.095711 | 0.124831 | 0.070845 | 0.06999 |
| ACECough | 0.562698 | 0.605251 | 0.201901 | 0.255772 | 0.039384 | 0.033952 |
| Diverticulosis | NA | NA | NA | NA | NA | NA |
| AAA | 0.986111 | 1 | 0.611111 | 0.611111 | 0.076389 | 0.097222 |

**Supplementary Table 13.** *Hits@100%* of individual phenotypes based on five seed concepts for EHR-based MCEs

|  | GloVeEmb_V | GloVeEmb_5Y | SGEmb_V | SGEmb_5Y |
| --- | --- | --- | --- | --- |
| VTE | 0.356082 | 0.439657 | 0.19168 | 0.140481 |
| Dementia | 0.096408 | 0.232514 | 0.141777 | 0.10775 |
| Zoster | 0.244063 | 0.191489 | 0.108814 | 0.059048 |
| DHaCKD | 0.221893 | 0.321006 | 0.177515 | 0.147929 |
| EO | 0.359088 | 0.378569 | 0.28388 | 0.332153 |
| CF | 0.319444 | 0.463333 | 0.340556 | 0.156389 |
| Height | 0.111531 | 0.081888 | 0.082398 | 0.051939 |
| Asthma | 0.124662 | 0.161603 | 0.14593 | 0.082272 |
| HF | 0.278754 | 0.367296 | 0.265734 | 0.150499 |
| CaMRSA | 0.120526 | 0.089116 | 0.065011 | 0.043828 |
| Hypothyroidism | 0.125732 | 0.152344 | 0.092529 | 0.053955 |
| Lipids | 0.340697 | 0.209003 | 0.188108 | 0.096957 |
| ECO | 0.071217 | 0.088417 | 0.05778 | 0.062618 |
| Autism | 0.417056 | 0.447205 | 0.21445 | 0.089635 |
| Satin/MACE | 0.222222 | 0.305556 | 0.166667 | 0.104167 |
| ADHD | 0.153653 | 0.2388 | 0.160313 | 0.103534 |
| RBC | 0.274447 | 0.210787 | 0.223123 | 0.121708 |
| AD | 0.20762 | 0.205787 | 0.117148 | 0.072165 |
| T2D | 0.11819 | 0.132656 | 0.092028 | 0.049246 |
| BPH | 0.201389 | 0.253472 | 0.076389 | 0.111111 |
| GERD | 0.121016 | 0.116624 | 0.109531 | 0.058871 |
| Appendicitis | 0.092349 | 0.092349 | 0.102309 | 0.061566 |
| Epilepsy | 0.56213 | 0.556213 | 0.207101 | 0.112426 |
| CRC | 0.352621 | 0.248798 | 0.23893 | 0.151059 |
| CIN | 0.308264 | 0.350773 | 0.228002 | 0.102854 |
| CR | 0.20734 | 0.320741 | 0.171 | 0.071531 |
| CKD | 0.210625 | 0.248572 | 0.141162 | 0.121508 |
| ResHTN | 0.091029 | 0.073061 | 0.075043 | 0.058925 |
| Cdiff | 0.272389 | 0.205901 | 0.126956 | 0.055178 |
| QRS | 0.359459 | 0.394439 | 0.33667 | 0.246298 |
| ACECough | 0.151701 | 0.19518 | 0.115784 | 0.07656 |
| Diverticulosis | NA | NA | NA | NA |
| AAA | 0.368056 | 0.541667 | 0.138889 | 0.131944 |

**Supplementary Table 14.** *Hits@200%* of individual phenotypes based on five seed concepts for graph-based MCEs

|  | n2vEmb | n2vEmb+ | LINEEmb | LINEEmb+ | SVDEmb | SVDEmb+ |
| --- | --- | --- | --- | --- | --- | --- |
| VTE | 0.640454 | 0.675272 | 0.298325 | 0.317412 | 0.306096 | 0.298014 |
| Dementia | 0.928819 | 0.928819 | 0.296875 | 0.420139 | 0.050347 | 0.072917 |
| Zoster | 0.920636 | 0.921855 | 0.359168 | 0.7366 | 0.102017 | 0.102064 |
| DHaCKD | 0.427515 | 0.486686 | 0.275148 | 0.488166 | 0.093195 | 0.059172 |
| EO | 0.7563 | 0.763555 | 0.599548 | 0.626428 | 0.622232 | 0.623656 |
| CF | 0.609977 | 0.685311 | 0.193752 | 0.374402 | 0.041824 | 0.039305 |
| Height | 0.552118 | 0.595842 | 0.141983 | 0.327633 | 0.082873 | 0.077125 |
| Asthma | 0.556667 | 0.585111 | 0.115111 | 0.146622 | 0.07 | 0.067289 |
| HF | 0.503471 | 0.540406 | 0.165717 | 0.323603 | 0.059986 | 0.054201 |
| CaMRSA | 0.508642 | 0.502716 | 0.220741 | 0.230123 | 0.054321 | 0.038025 |
| Hypothyroidism | 0.579022 | 0.637689 | 0.149511 | 0.232356 | 0.034667 | 0.04 |
| Lipids | 0.912639 | 0.919609 | 0.294492 | 0.426457 | 0.240925 | 0.260351 |
| ECO | 0.390023 | 0.459814 | 0.226002 | 0.290753 | 0.044848 | 0.047619 |
| Autism | 0.891029 | 0.945076 | 0.199687 | 0.245666 | 0.159648 | 0.16152 |
| Satin/MACE | 0.852071 | 0.893491 | 0.64497 | 0.852071 | 0.094675 | 0.094675 |
| ADHD | 0.703271 | 0.753674 | 0.224596 | 0.299259 | 0.208069 | 0.210913 |
| RBC | 0.598584 | 0.614614 | 0.308104 | 0.426448 | 0.247651 | 0.255466 |
| AD | 0.492925 | 0.516465 | 0.135324 | 0.231688 | 0.088043 | 0.080211 |
| T2D | 0.480516 | 0.489385 | 0.19269 | 0.263101 | 0.051599 | 0.04058 |
| BPH | 0.675781 | 0.796875 | 0.199219 | 0.27832 | 0.043945 | 0.068359 |
| GERD | 0.842784 | 0.844038 | 0.149273 | 0.351668 | 0.074472 | 0.082206 |
| Appendicitis | 0.3852 | 0.4568 | 0.14 | 0.1704 | 0.042 | 0.0444 |
| Epilepsy | 1 | 1 | 0.47929 | 0.579882 | 0.076923 | 0.106509 |
| CRC | 0.951826 | 0.957939 | 0.337455 | 0.420424 | 0.336754 | 0.342008 |
| CIN | 0.368572 | 0.359092 | 0.183855 | 0.370009 | 0.047113 | 0.037346 |
| CR | 0.802336 | 0.842565 | 0.144457 | 0.214125 | 0.056622 | 0.062564 |
| CKD | 0.913117 | 0.919762 | 0.68623 | 0.715528 | 0.545498 | 0.53833 |
| ResHTN | 0.355651 | 0.424076 | 0.131948 | 0.208607 | 0.048231 | 0.051858 |
| Cdiff | 0.793234 | 0.788716 | 0.135212 | 0.225779 | 0.099923 | 0.101787 |
| QRS | 0.624984 | 0.639083 | 0.161617 | 0.195361 | 0.136929 | 0.139326 |
| ACECough | 0.770937 | 0.812585 | 0.245813 | 0.301041 | 0.044364 | 0.039837 |
| Diverticulosis | NA | NA | NA | NA | NA | NA |
| AAA | 1 | 1 | 0.75 | 0.673611 | 0.083333 | 0.111111 |

**Supplementary Table 15.** *Hits@200%* of individual phenotypes based on five seed concepts for EHR-based MCEs

|  | GloVeEmb_V | GloVeEmb_5Y | SGEmb_V | SGEmb_5Y |
| --- | --- | --- | --- | --- |
| VTE | 0.491389 | 0.570064 | 0.293649 | 0.30925 |
| Dementia | 0.126654 | 0.327032 | 0.219282 | 0.117202 |
| Zoster | 0.349332 | 0.251301 | 0.158406 | 0.093651 |
| DHaCKD | 0.295858 | 0.408284 | 0.263314 | 0.171598 |
| EO | 0.683939 | 0.695144 | 0.646075 | 0.690827 |
| CF | 0.454444 | 0.630278 | 0.471111 | 0.265 |
| Height | 0.171173 | 0.150918 | 0.137653 | 0.083316 |
| Asthma | 0.240066 | 0.270014 | 0.246335 | 0.147473 |
| HF | 0.42947 | 0.539822 | 0.412218 | 0.26378 |
| CaMRSA | 0.156318 | 0.135135 | 0.091308 | 0.055515 |
| Hypothyroidism | 0.172852 | 0.223633 | 0.138184 | 0.0896 |
| Lipids | 0.496849 | 0.343361 | 0.316573 | 0.210111 |
| ECO | 0.107498 | 0.13061 | 0.082505 | 0.074174 |
| Autism | 0.542763 | 0.566419 | 0.303828 | 0.192834 |
| Satin/MACE | 0.270833 | 0.340278 | 0.229167 | 0.104167 |
| ADHD | 0.291143 | 0.3885 | 0.292493 | 0.227773 |
| RBC | 0.442389 | 0.364721 | 0.354181 | 0.25406 |
| AD | 0.29873 | 0.31996 | 0.189765 | 0.137103 |
| T2D | 0.164666 | 0.202524 | 0.129578 | 0.078178 |
| BPH | 0.248264 | 0.295139 | 0.098958 | 0.119792 |
| GERD | 0.177773 | 0.185872 | 0.158438 | 0.100845 |
| Appendicitis | 0.135355 | 0.144409 | 0.146673 | 0.074694 |
| Epilepsy | 0.656805 | 0.751479 | 0.236686 | 0.118343 |
| CRC | 0.549389 | 0.475037 | 0.400236 | 0.337277 |
| CIN | 0.454816 | 0.494649 | 0.318668 | 0.19679 |
| CR | 0.303577 | 0.450589 | 0.249066 | 0.160658 |
| CKD | 0.349355 | 0.390141 | 0.244867 | 0.205534 |
| ResHTN | 0.149161 | 0.128286 | 0.124455 | 0.099221 |
| Cdiff | 0.408471 | 0.30016 | 0.185661 | 0.091683 |
| QRS | 0.551479 | 0.586459 | 0.469956 | 0.375309 |
| ACECough | 0.230624 | 0.285917 | 0.157372 | 0.110586 |
| Diverticulosis | NA | NA | NA | NA |
| AAA | 0.388889 | 0.590278 | 0.236111 | 0.131944 |

**Supplementary Table 16.** *Hits@500%* of individual phenotypes based on five seed concepts for graph-based MCEs

|  | n2vEmb | n2vEmb+ | LINEEmb | LINEEmb+ | SVDEmb | SVDEmb+ |
| --- | --- | --- | --- | --- | --- | --- |
| VTE | 0.896811 | 0.912711 | 0.74574 | 0.747396 | 0.758712 | 0.753298 |
| Dementia | 0.987847 | 0.979167 | 0.350694 | 0.46875 | 0.057292 | 0.090278 |
| Zoster | 0.945492 | 0.941165 | 0.559156 | 0.857879 | 0.25398 | 0.260635 |
| DHaCKD | 0.609467 | 0.643491 | 0.329882 | 0.579882 | 0.122781 | 0.076923 |
| EO | 1 | 1 | 1 | 1 | 1 | 1 |
| CF | 0.796674 | 0.817334 | 0.264046 | 0.456538 | 0.085664 | 0.079365 |
| Height | 0.664646 | 0.716864 | 0.261546 | 0.433956 | 0.191941 | 0.185993 |
| Asthma | 0.712711 | 0.739289 | 0.202089 | 0.242489 | 0.160667 | 0.159422 |
| HF | 0.733446 | 0.737095 | 0.252581 | 0.433695 | 0.118014 | 0.126023 |
| CaMRSA | 0.678519 | 0.671111 | 0.308642 | 0.299259 | 0.073086 | 0.082469 |
| Hypothyroidism | 0.654578 | 0.730133 | 0.215822 | 0.316267 | 0.071289 | 0.087289 |
| Lipids | 0.942981 | 0.948736 | 0.6181 | 0.686781 | 0.612357 | 0.630483 |
| ECO | 0.539934 | 0.593348 | 0.317712 | 0.398841 | 0.07357 | 0.075082 |
| Autism | 0.9594 | 0.984808 | 0.439985 | 0.495325 | 0.391035 | 0.385413 |
| Satin/MACE | 0.911243 | 0.940828 | 0.721893 | 0.887574 | 0.100592 | 0.118343 |
| ADHD | 0.878653 | 0.919902 | 0.527183 | 0.592148 | 0.519035 | 0.526007 |
| RBC | 0.840523 | 0.856748 | 0.643105 | 0.715163 | 0.622793 | 0.624026 |
| AD | 0.646872 | 0.672326 | 0.256185 | 0.368525 | 0.208593 | 0.197935 |
| T2D | 0.604407 | 0.634776 | 0.26122 | 0.339962 | 0.086536 | 0.076055 |
| BPH | 0.859375 | 0.929688 | 0.24707 | 0.34668 | 0.05957 | 0.092773 |
| GERD | 0.874287 | 0.885395 | 0.277285 | 0.508227 | 0.181522 | 0.195258 |
| Appendicitis | 0.5336 | 0.6052 | 0.1968 | 0.2188 | 0.0664 | 0.0688 |
| Epilepsy | 1 | 1 | 0.544379 | 0.639053 | 0.076923 | 0.106509 |
| CRC | 0.984996 | 0.98551 | 0.81244 | 0.837201 | 0.837644 | 0.83197 |
| CIN | 0.561333 | 0.59006 | 0.238725 | 0.4447 | 0.085033 | 0.08216 |
| CR | 0.891196 | 0.934909 | 0.237962 | 0.32409 | 0.123967 | 0.136603 |
| CKD | 1 | 1 | 1 | 1 | 1 | 1 |
| ResHTN | 0.489266 | 0.55769 | 0.204 | 0.289383 | 0.099402 | 0.119596 |
| Cdiff | 0.991535 | 0.992878 | 0.262094 | 0.356658 | 0.24392 | 0.245297 |
| QRS | 0.80769 | 0.813719 | 0.346631 | 0.385355 | 0.337471 | 0.365945 |
| ACECough | 0.940697 | 0.939792 | 0.30919 | 0.358533 | 0.076053 | 0.070167 |
| Diverticulosis | NA | NA | NA | NA | NA | NA |
| AAA | 1 | 1 | 0.798611 | 0.729167 | 0.083333 | 0.118056 |

**Supplementary Table 17.** *Hits@500%* of individual phenotypes based on five seed concepts for EHR-based MCEs

|  | GloVeEmb_V | GloVeEmb_5Y | SGEmb_V | SGEmb_5Y |
| --- | --- | --- | --- | --- |
| VTE | 0.895468 | 0.944618 | 0.848208 | 0.871378 |
| Dementia | 0.266541 | 0.463138 | 0.311909 | 0.130435 |
| Zoster | 0.493502 | 0.376047 | 0.252097 | 0.212115 |
| DHaCKD | 0.423077 | 0.600592 | 0.420118 | 0.251479 |
| EO | 1 | 1 | 1 | 1 |
| CF | 0.712222 | 0.779444 | 0.656944 | 0.410556 |
| Height | 0.301633 | 0.295408 | 0.25051 | 0.201276 |
| Asthma | 0.485147 | 0.496383 | 0.432292 | 0.305845 |
| HF | 0.670139 | 0.78418 | 0.584527 | 0.447049 |
| CaMRSA | 0.241052 | 0.251278 | 0.147553 | 0.129291 |
| Hypothyroidism | 0.248535 | 0.384521 | 0.24585 | 0.172363 |
| Lipids | 0.776057 | 0.737769 | 0.634622 | 0.608721 |
| ECO | 0.187853 | 0.231389 | 0.13921 | 0.110723 |
| Autism | 0.756472 | 0.779174 | 0.579639 | 0.502398 |
| Satin/MACE | 0.388889 | 0.513889 | 0.284722 | 0.159722 |
| ADHD | 0.700829 | 0.748709 | 0.640514 | 0.642002 |
| RBC | 0.758403 | 0.75833 | 0.684817 | 0.686542 |
| AD | 0.497742 | 0.547375 | 0.356972 | 0.308827 |
| T2D | 0.270545 | 0.337335 | 0.198215 | 0.192367 |
| BPH | 0.329861 | 0.354167 | 0.140625 | 0.180556 |
| GERD | 0.319062 | 0.346129 | 0.269844 | 0.212453 |
| Appendicitis | 0.225441 | 0.261204 | 0.225441 | 0.155274 |
| Epilepsy | 0.692308 | 0.846154 | 0.35503 | 0.136095 |
| CRC | 0.958032 | 0.970582 | 0.948052 | 0.937804 |
| CIN | 0.631688 | 0.718787 | 0.465517 | 0.492271 |
| CR | 0.486354 | 0.64098 | 0.379704 | 0.297831 |
| CKD | 0.619283 | 0.690849 | 0.494072 | 0.488546 |
| ResHTN | 0.327124 | 0.288281 | 0.238473 | 0.233849 |
| Cdiff | 0.562274 | 0.491868 | 0.319728 | 0.213096 |
| QRS | 0.837621 | 0.883126 | 0.784451 | 0.692872 |
| ACECough | 0.35775 | 0.466446 | 0.252836 | 0.264178 |
| Diverticulosis | NA | NA | NA | NA |
| AAA | 0.430556 | 0.645833 | 0.361111 | 0.152778 |

**Supplementary Table 18.** *Hits@1000%* of individual phenotypes based on five seed concepts for graph-based MCEs

|  | n2vEmb | n2vEmb+ | LINEEmb | LINEEmb+ | SVDEmb | SVDEmb+ |
| --- | --- | --- | --- | --- | --- | --- |
| VTE | 1 | 1 | 1 | 1 | 1 | 1 |
| Dementia | 0.998264 | 0.993056 | 0.423611 | 0.510417 | 0.065972 | 0.104167 |
| Zoster | 0.972535 | 0.96727 | 0.770704 | 0.942586 | 0.507476 | 0.506116 |
| DHaCKD | 0.776627 | 0.810651 | 0.380178 | 0.647929 | 0.153846 | 0.113905 |
| EO | 1 | 1 | 1 | 1 | 1 | 1 |
| CF | 0.877299 | 0.886369 | 0.334089 | 0.534643 | 0.149408 | 0.141345 |
| Height | 0.760645 | 0.81558 | 0.44568 | 0.575853 | 0.367954 | 0.378764 |
| Asthma | 0.829244 | 0.842133 | 0.333867 | 0.382356 | 0.304178 | 0.2944 |
| HF | 0.852083 | 0.86303 | 0.374155 | 0.54717 | 0.214756 | 0.26157 |
| CaMRSA | 0.785679 | 0.773827 | 0.387654 | 0.378272 | 0.113086 | 0.142222 |
| Hypothyroidism | 0.712 | 0.793956 | 0.293156 | 0.402844 | 0.142756 | 0.157689 |
| Lipids | 1 | 1 | 1 | 1 | 1 | 1 |
| ECO | 0.654069 | 0.687075 | 0.414462 | 0.502394 | 0.129504 | 0.141849 |
| Autism | 0.994973 | 0.998613 | 0.823109 | 0.854481 | 0.800799 | 0.782377 |
| Satin/MACE | 0.982249 | 0.952663 | 0.757396 | 0.905325 | 0.100592 | 0.136095 |
| ADHD | 1 | 1 | 1 | 1 | 1 | 1 |
| RBC | 1 | 1 | 1 | 1 | 1 | 1 |
| AD | 0.778992 | 0.806537 | 0.438212 | 0.547593 | 0.409154 | 0.380785 |
| T2D | 0.708143 | 0.733405 | 0.337812 | 0.419242 | 0.129535 | 0.140285 |
| BPH | 0.94043 | 0.980469 | 0.291016 | 0.404297 | 0.095703 | 0.128906 |
| GERD | 0.903939 | 0.915793 | 0.464929 | 0.677715 | 0.349697 | 0.371973 |
| Appendicitis | 0.644 | 0.6872 | 0.256 | 0.2768 | 0.108 | 0.1236 |
| Epilepsy | 1 | 1 | 0.60355 | 0.710059 | 0.112426 | 0.106509 |
| CRC | 1 | 1 | 1 | 1 | 1 | 1 |
| CIN | 0.688308 | 0.733984 | 0.300776 | 0.507038 | 0.12956 | 0.150244 |
| CR | 0.929991 | 0.973226 | 0.373608 | 0.466088 | 0.247183 | 0.253125 |
| CKD | 1 | 1 | 1 | 1 | 1 | 1 |
| ResHTN | 0.660229 | 0.703362 | 0.302813 | 0.390844 | 0.183904 | 0.224684 |
| Cdiff | 0.996322 | 0.997279 | 0.457908 | 0.532904 | 0.479928 | 0.488108 |
| QRS | 0.946757 | 0.944296 | 0.658857 | 0.692342 | 0.675329 | 0.704077 |
| ACECough | 0.958805 | 0.966048 | 0.382526 | 0.422363 | 0.117248 | 0.129018 |
| Diverticulosis | NA | NA | NA | NA | NA | NA |
| AAA | 1 | 1 | 0.861111 | 0.791667 | 0.138889 | 0.125 |

**Supplementary Table 19.** *Hits@1000%* of individual phenotypes based on five seed concepts for EHR-based MCEs

|  | GloVeEmb_V | GloVeEmb_5Y | SGEmb_V | SGEmb_5Y |
| --- | --- | --- | --- | --- |
| VTE | 1 | 1 | 1 | 1 |
| Dementia | 0.417769 | 0.542533 | 0.340265 | 0.162571 |
| Zoster | 0.647921 | 0.543261 | 0.403414 | 0.459286 |
| DHaCKD | 0.664201 | 0.748521 | 0.597633 | 0.390533 |
| EO | 1 | 1 | 1 | 1 |
| CF | 0.838611 | 0.921111 | 0.781111 | 0.579167 |
| Height | 0.469541 | 0.487806 | 0.449133 | 0.367602 |
| Asthma | 0.711034 | 0.742814 | 0.664159 | 0.508632 |
| HF | 0.815647 | 0.932834 | 0.70931 | 0.659288 |
| CaMRSA | 0.332359 | 0.439737 | 0.248356 | 0.216947 |
| Hypothyroidism | 0.328369 | 0.58374 | 0.320801 | 0.30835 |
| Lipids | 1 | 1 | 1 | 1 |
| ECO | 0.28702 | 0.382693 | 0.217146 | 0.215802 |
| Autism | 1 | 1 | 1 | 1 |
| Satin/MACE | 0.458333 | 0.722222 | 0.444444 | 0.208333 |
| ADHD | 1 | 1 | 1 | 1 |
| RBC | 1 | 1 | 1 | 1 |
| AD | 0.748114 | 0.778776 | 0.647997 | 0.591827 |
| T2D | 0.389966 | 0.484149 | 0.256694 | 0.344721 |
| BPH | 0.416667 | 0.416667 | 0.182292 | 0.236111 |
| GERD | 0.549844 | 0.540951 | 0.469844 | 0.429112 |
| Appendicitis | 0.359891 | 0.40833 | 0.340878 | 0.297872 |
| Epilepsy | 0.715976 | 0.863905 | 0.443787 | 0.153846 |
| CRC | 1 | 1 | 1 | 1 |
| CIN | 0.739298 | 0.840963 | 0.596908 | 0.612663 |
| CR | 0.656421 | 0.788136 | 0.557383 | 0.506679 |
| CKD | 0.983868 | 0.990832 | 0.988584 | 0.985815 |
| ResHTN | 0.541815 | 0.496103 | 0.389748 | 0.386313 |
| Cdiff | 0.741105 | 0.728423 | 0.522354 | 0.458666 |
| QRS | 0.982758 | 0.98719 | 0.9831 | 0.960567 |
| ACECough | 0.507561 | 0.624764 | 0.356333 | 0.30482 |
| Diverticulosis | NA | NA | NA | NA |
| AAA | 0.513889 | 0.791667 | 0.444444 | 0.166667 |

**Supplementary Table 20.** *Hits@2000%* of individual phenotypes based on five seed concepts for graph-based MCEs

|  | n2vEmb | n2vEmb+ | LINEEmb | LINEEmb+ | SVDEmb | SVDEmb+ |
| --- | --- | --- | --- | --- | --- | --- |
| VTE | 1 | 1 | 1 | 1 | 1 | 1 |
| Dementia | 1 | 1 | 0.510417 | 0.581597 | 0.112847 | 0.147569 |
| Zoster | 0.999969 | 0.999703 | 0.995157 | 0.998547 | 0.975753 | 0.975597 |
| DHaCKD | 0.828402 | 0.85503 | 0.449704 | 0.70858 | 0.193787 | 0.189349 |
| EO | 1 | 1 | 1 | 1 | 1 | 1 |
| CF | 0.93424 | 0.946082 | 0.444444 | 0.639204 | 0.275888 | 0.28773 |
| Height | 0.888759 | 0.936201 | 0.771455 | 0.818668 | 0.706883 | 0.733249 |
| Asthma | 0.945467 | 0.938933 | 0.588311 | 0.629733 | 0.586978 | 0.544667 |
| HF | 0.942951 | 0.949448 | 0.563724 | 0.707191 | 0.393289 | 0.481132 |
| CaMRSA | 0.875556 | 0.859753 | 0.495309 | 0.484444 | 0.204444 | 0.265185 |
| Hypothyroidism | 0.778311 | 0.863289 | 0.424178 | 0.5344 | 0.288533 | 0.300089 |
| Lipids | 1 | 1 | 1 | 1 | 1 | 1 |
| ECO | 0.771983 | 0.78584 | 0.54069 | 0.628874 | 0.244142 | 0.250441 |
| Autism | 1 | 1 | 1 | 1 | 1 | 1 |
| Satin/MACE | 1 | 0.952663 | 0.798817 | 0.928994 | 0.106509 | 0.183432 |
| ADHD | 1 | 1 | 1 | 1 | 1 | 1 |
| RBC | 1 | 1 | 1 | 1 | 1 | 1 |
| AD | 0.938012 | 0.957614 | 0.805669 | 0.848745 | 0.807672 | 0.813991 |
| T2D | 0.803547 | 0.821016 | 0.458479 | 0.535609 | 0.241602 | 0.27197 |
| BPH | 0.980469 | 1 | 0.375977 | 0.482422 | 0.166016 | 0.180664 |
| GERD | 0.959808 | 0.95936 | 0.791006 | 0.889516 | 0.68312 | 0.686464 |
| Appendicitis | 0.7408 | 0.7584 | 0.35 | 0.372 | 0.2096 | 0.2252 |
| Epilepsy | 1 | 1 | 0.686391 | 0.745562 | 0.195266 | 0.12426 |
| CRC | 1 | 1 | 1 | 1 | 1 | 1 |
| CIN | 0.812698 | 0.832232 | 0.395289 | 0.610457 | 0.209135 | 0.305085 |
| CR | 0.962298 | 0.98996 | 0.609385 | 0.678437 | 0.490609 | 0.467045 |
| CKD | 1 | 1 | 1 | 1 | 1 | 1 |
| ResHTN | 0.860504 | 0.879129 | 0.473287 | 0.564651 | 0.382904 | 0.416822 |
| Cdiff | 0.99995 | 1 | 0.912658 | 0.917075 | 0.93708 | 0.935602 |
| QRS | 1 | 1 | 1 | 1 | 1 | 1 |
| ACECough | 0.966501 | 0.971933 | 0.481213 | 0.50928 | 0.187415 | 0.230874 |
| Diverticulosis | NA | NA | NA | NA | NA | NA |
| AAA | 1 | 1 | 0.881944 | 0.819444 | 0.138889 | 0.138889 |

**Supplementary Table 21.** *Hits@2000%* of individual phenotypes based on five seed concepts for EHR-based MCEs

|  | GloVeEmb_V | GloVeEmb_5Y | SGEmb_V | SGEmb_5Y |
| --- | --- | --- | --- | --- |
| VTE | 1 | 1 | 1 | 1 |
| Dementia | 0.478261 | 0.674858 | 0.412098 | 0.294896 |
| Zoster | 1 | 1 | 1 | 1 |
| DHaCKD | 0.826923 | 0.884615 | 0.70858 | 0.585799 |
| EO | 1 | 1 | 1 | 1 |
| CF | 0.952222 | 0.994167 | 0.878889 | 0.760278 |
| Height | 0.729541 | 0.787959 | 0.738724 | 0.733214 |
| Asthma | 0.934462 | 0.967062 | 0.917245 | 0.855228 |
| HF | 0.963325 | 0.989583 | 0.872504 | 0.797526 |
| CaMRSA | 0.483565 | 0.664719 | 0.426589 | 0.472608 |
| Hypothyroidism | 0.457764 | 0.781494 | 0.423828 | 0.490967 |
| Lipids | 1 | 1 | 1 | 1 |
| ECO | 0.453641 | 0.596345 | 0.417092 | 0.40473 |
| Autism | 1 | 1 | 1 | 1 |
| Satin/MACE | 0.597222 | 0.847222 | 0.5625 | 0.298611 |
| ADHD | 1 | 1 | 1 | 1 |
| RBC | 1 | 1 | 1 | 1 |
| AD | 1 | 1 | 1 | 1 |
| T2D | 0.59526 | 0.675592 | 0.389966 | 0.550939 |
| BPH | 0.482639 | 0.494792 | 0.258681 | 0.274306 |
| GERD | 0.846602 | 0.906717 | 0.854102 | 0.882103 |
| Appendicitis | 0.556813 | 0.6555 | 0.533273 | 0.535084 |
| Epilepsy | 0.786982 | 0.905325 | 0.47929 | 0.213018 |
| CRC | 1 | 1 | 1 | 1 |
| CIN | 0.838585 | 0.908145 | 0.770511 | 0.770511 |
| CR | 0.835464 | 0.877837 | 0.814278 | 0.768601 |
| CKD | 1 | 1 | 1 | 1 |
| ResHTN | 0.794028 | 0.810807 | 0.705113 | 0.655305 |
| Cdiff | 1 | 1 | 1 | 1 |
| QRS | 1 | 1 | 1 | 1 |
| ACECough | 0.64414 | 0.751418 | 0.55104 | 0.572779 |
| Diverticulosis | NA | NA | NA | NA |
| AAA | 0.576389 | 0.881944 | 0.520833 | 0.236111 |
